# The Potential Clinical Impact of Implementing Different COVID-19 Boosters in Fall 2022 in the United States

**DOI:** 10.1101/2022.07.19.22277824

**Authors:** Michele A. Kohli, Michael Maschio, Amy Lee, Kelly Fust, Nicolas Van de Velde, Philip O. Buck, Milton C. Weinstein

## Abstract

**Objective:** Emerging SARS-COV-2 variants are spurring the development of adapted vaccines as public health authorities plan for the fall vaccination strategy. We aimed to estimate the number of infections and hospitalizations prevented by three potential booster strategies in those ≥18 years of age in the United States: Boosting with Moderna’s licensed first generation monovalent vaccine mRNA-1273 (ancestral strain) starting in September 2022, boosting with Moderna’s candidate bivalent vaccine mRNA-1273.214 (ancestral + BA.1 variant of concern [VOC]) starting in September 2022, or boosting with Moderna’s updated candidate bivalent vaccine mRNA-1273.222 (ancestral + BA.4/5 VOC) starting 2 months later in November 2022 due to longer development timeline.

**Methods:** An age-stratified, transmission dynamic, Susceptible-Exposed-Infection-Recovered (SEIR) model, adapted from previous literature, was used to estimate the number of infections over time; the model contains compartments defined by both SEIR status and vaccination status. A decision tree was subsequently used to estimate the clinical consequences of those infections. Calibration was performed so the model tracks the actual course of the pandemic up to the present time.

**Results:** Vaccinating with mRNA-1273(Sept), mRNA-1273.214(Sept), and mRNA-1273.222(Nov) is predicted to reduce infections by 34%, 40%, and 18%, respectively, over a 6-month time horizon (September-February) compared to no booster. Similarly, boosting in September prevents substantially more hospitalizations than starting to boost in November with a more effective vaccine (42%, 48%, and 25% for mRNA-1273, mRNA-1273.214, and mRNA-1273.222, respectively, at 6 months compared to no booster). Sensitivity analyses around transmissibility, vaccine coverage, masking, and waning of natural and vaccine-induced immunity changed the magnitude of cases prevented but boosting with mRNA-1273.214 in September consistently prevented more cases of infection and hospitalization than the other two strategies.

**Limitations and Conclusions:** With the emergence of new variants, key characteristics of the virus that affect estimates of spread and clinical impact also evolve, making estimation of these parameters difficult, especially in heterogeneous populations. Our analysis demonstrated that vaccinating with the bivalent mRNA-1273.214 booster was more effective over a 6-month period in preventing infections and hospitalizations with a BA.4/5 subvariant than the tailored vaccine, simply because it could be deployed 2 months earlier. We conclude that there is no advantage to delay boosting until a BA.4/5 vaccine is available; earlier boosting with mRNA-1273.214 will prevent the most infections and hospitalizations.

## 1. INTRODUCTION

The COVID-19 pandemic has been marked by the emergence of multiple variants of concern (VOC), including most recently Omicron, which arose in October 2021 and rapidly replaced all other circulating variants globally.^1^ The World Health Organization (WHO) has stated that the available evidence suggests that Omicron VOC is more transmissible and shows evidence of immune escape, as there is a reduction in neutralizing activity of antibody responses compared to previous VOCs.^2^ The original Omicron VOC, now termed BA.1, has already evolved into new sublineages. In the United States (US), the proportion of infections attributed to the BA.4 and BA.5 sublineages is increasing and, as of July 6, 2022, these are predicted to be the most common sublineages in the near future.^3,4^ While the characteristics of BA.4 and BA.5 are still being evaluated relative to BA.1, one concerning observation is that individuals produce lower neutralizing antibody titers compared to BA.1.^2^

Since the successful launch of vaccines designed to protect against ancestral SARS-COV-2 infections, vaccine manufacturers have been developing new adapted vaccines targeting emerging VOCs. In April, Moderna reported superior neutralizing antibody responses generated by its candidate mRNA-1273.211 bivalent booster that encodes for the spike glycoproteins of the ancestral strain and Beta (B.1.351) variant.^5^ On June 8, 2022, Moderna reported superior antibody responses for its mRNA-1273.214 bivalent booster, that encodes for the ancestral strain and Omicron (BA.1) VOC.^6,7^ Data from this latest clinical trial have now been submitted for review with regulators. In addition to superior neutralizing antibody responses to Omicron, next generation bivalent vaccines have also demonstrated a broader response to other VOCs, and this response was more durable for mRNA-1273.211 compared to the first generation monovalent vaccine, mRNA-1273.^4^

In anticipation of the peak winter season for respiratory viruses in the Northern Hemisphere and the registration of next generation COVID-19 vaccines, the WHO, European Medicines Agency (EMA), and US Food and Drug Administration (FDA) held several meetings where current epidemiological and vaccines data were reviewed. The WHO Technical Advisory Group on COVID-19 Vaccine Composition (TAG-CO-VAC) released an new interim statement in June.^1^ In this statement, TAG-CO-VAC reiterated that the primary objective of COVID-19 vaccination is to reduce hospitalization, severe disease and death, against which currently licensed vaccines continue to provide high levels of protection. However, in the context of uncertainty about the timing of the emergence, extent of global circulation, and antigenic characteristics of future variants, TAG-CO-VAC recommended that an additional vaccination objective be to broaden the immune response against circulating and emerging variants. As the most antigenically distinct SARS-CoV-2 VOC, they concluded that adding Omicron in an updated vaccine composition that includes the ancestral strain may be beneficial if administered as a booster dose. In their press briefing of July 7, 2022, EMA also communicated that the choice of vaccines will be based on the breadth of immunity against VOCs, given that no one can predict what variants will be circulating during the winter.^8^ Finally, following the Vaccines and Related Biological Products Advisory Committee (VRBPAC) vote on June 28, 2022, the FDA recommended for vaccine manufacturers to update booster formulations to include the ancestral strain and Omicron BA.4/5 given its current prevalence in the US.^9^ Such an update from the Omicron BA.1 currently included in candidate vaccines to BA.4/5 may have implications in terms of the timing of the fall vaccination strategy.

Our research objective was to estimate the number of infections and hospitalizations prevented by a booster strategy in those 18 years of age and older in the United States across a 6-month time horizon. We compared the following three scenarios: (1) Boosting with Moderna’s licensed first generation monovalent vaccine mRNA-1273 (ancestral) starting in September 2022, (2) Boosting with Moderna’s candidate bivalent vaccine mRNA-1273.214 (ancestral + BA.1 VOC) starting in September 2022, or (3) Boosting with Moderna’s updated candidate bivalent vaccine mRNA-1273.222 (ancestral + BA.4/5 VOC) starting in November 2022 due to longer development time.

## 2. METHODS

For this analysis, we use a simple, age-stratified, Susceptible-Exposed-Infection-Recovered (SEIR) compartmental model, adapted from Shiri et al., 2021^10^ to estimate the number of infections over time and then use a decision tree to estimate the clinical consequences of those infections across a 6-month time horizon (September 2022 to February 2023), where healthcare systems are the most pressured. The model structure, inputs, and calibration process are described in more detail in the accompanying technical appendix; a brief overview of key features and inputs is presented here.

### SEIR Model Structure

The SEIR model contains compartments defined by both SEIR status and vaccination status. The time step for transitions in the model is one day. The model simulation begins on January 31, 2020 and, as described in more detail below; calibration methods are used to ensure that the model tracks as well as possible the actual course of the pandemic up to the present time, but prior to implementation of a new booster vaccine. As shown in Figure 1, all individuals start in the unvaccinated compartments at the beginning of the pandemic. Individuals can move to the primary series compartments, then the booster 1 compartments, then the booster 2 compartments as more doses of vaccines are received. “Booster 1” and “booster 2” refer to doses of the currently available vaccine given after the primary series. For the primary series and each of these boosters, we assumed that a mix of the available vaccines (mRNA-1273; BNT162b2; AD26.COV2.S) were given based on US market shares at that time. We assume that no individuals receive primary series or booster 1 doses after May 31, 2022, and that no booster 2 doses are given after June 15, 2022.

**Figure 1.**
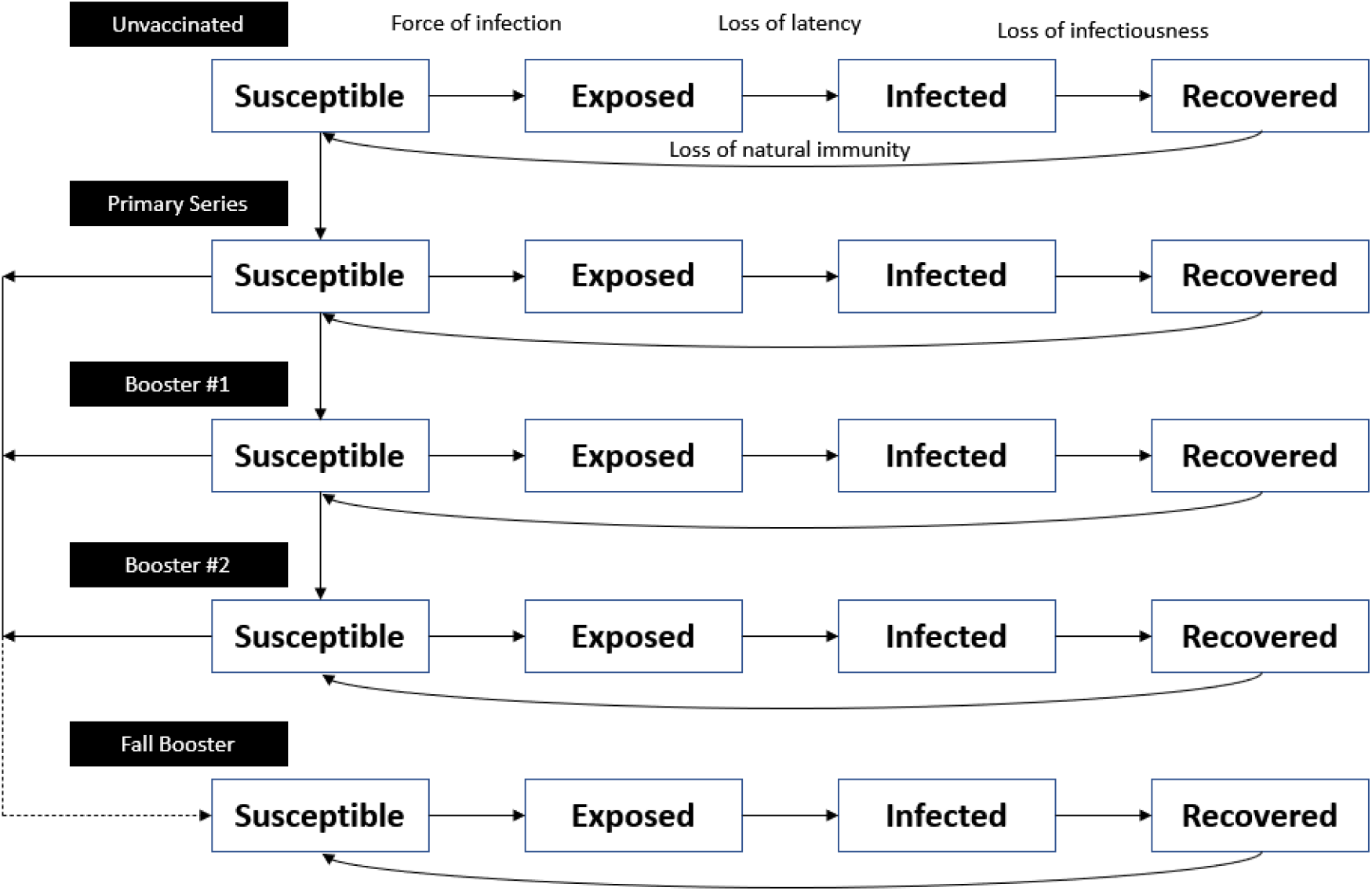
Model structure: Compartments in the dynamic model.

For this analysis, individuals may move into a final “Fall Booster” stratum when they receive a new booster starting in September or November 2022 (Figure 1). Individuals may receive this fall booster if they have received the primary series, booster 1, or booster 2.

All individuals start in the susceptible (S) compartments and move through to the recovered (R) compartments as they develop an infection followed by natural immunity. The force of infection, which is indicated by arrows in the model figure, is a function of effective contact between the susceptible and infected people. This rate is dictated by an age-specific contact matrix, which is modified by reduction in social mobility and masking behavior, and transmissibility of the virus. Vaccination also acts to reduce the force of infection. As described in the technical appendix, the model was calibrated between January 31, 2020 to May 31, 2022 to match all infections as estimated by the Institute for Health Metrics Evaluation (IHME).^11^ This process gave us an estimate of the number of people who had natural immunity and the average vaccine effectiveness (VE) for the proportions of the population in each of the vaccination strata, by age group in June 2022.

### Infection Projections

For projections beyond May 2022, it was assumed that social distancing patterns would return to pre-pandemic levels and mask use would return to 0% by the end of July 2022. Both remain at these pre-pandemic levels for the remainder of the analysis time horizon. Projections of the potential impact of a new variant similar to the BA.4/5 sub-variant are highly variable.^12^ Initial indications from countries such as Portugal that have experienced an early BA.4/5 wave^13^ are that BA.4/5 will not be as severe as the first BA.1 wave, but will be more severe than the BA.2 waves. We assumed that BA.4/5 would be the only sub-variants circulating by August 15, 2022, and therefore increased our transmissibility parameter until September 1, 2022, and held it constant thereafter. For the base case, we assumed that the peak incidence of infection without boosters would be approximately half of the Omicron BA.1 wave.

### Vaccine Effectiveness

In order to model the impact of a changing mix of VOC on VE, we divided the model simulation timeframe into three periods: pre-Omicron (January 31, 2020 – November 30, 2021); Omicron BA.1/2 (December 1, 2021 – August 14, 2022); and Omicron BA.4/5 (August 15, 2022 – February 28, 2023).

VE was estimated for primary series, booster 1, and booster 2 using a weighted average of the types of vaccines received in the US. For the pre-Omicron period, initial VE (against infection and severe disease) and the monthly waning rates were obtained from an analysis of 20 studies conducted by the IHME.^14^ Data for boosters were obtained from a test-negative case-control study conducted in England, with assumptions applied for missing data.^15^ For the Omicron BA.1 period, initial VE for primary series and booster and waning for the primary series were obtained from a meta-analysis by Pratama et al, 2022;^16^ data collected in England^17^ were used where meta-analytic data were unavailable. The waning rates for the booster were assumed to be the same as the waning rates for the primary series. The values used in the model are displayed in Table 1, while further details are presented in the technical appendix.

**Table 1.** Vaccine effectiveness for primary series, booster 1 and booster 2 (pre-Omicron and Omicron BA.1 periods)

| Vaccine | Pre-Omicron |  | Omicron (BA.1) |  |
| --- | --- | --- | --- | --- |
|  | Initial Vaccine Effectiveness | Waning: Monthly Decline | Initial Vaccine Effectiveness | Waning: Monthly Decline |
| Infections |  |  |  |  |
| Primary Series | 84.9% | 3.8% | 64.7% | 7.3% |
| Booster 1 | 93.2% | 3.7% | 72.9% | 7.3% |
| Booster 2 | NA |  | 72.9% | 7.3% |
| Severe Infections |  |  |  |  |
| Primary Series | 94.2% | 1.3% | 88.2% | 3.4% |
| Booster 1 | 99.7% | 1.2% | 93.3% | 3.4% |
| Booster 2 | NA |  | 93.3% | 3.4% |

Real-world VE data against the BA.4/5 variant are not yet available. However, the method developed by Khoury et al., 2021,^18^ and updated by Hogan et al., 2021,^19^ which calculates VE over time based on relative neutralizing antibody levels (geometric mean titers, GMT), was used to estimate the impact of new vaccines and emerging variants. Moderna clinical data showed a 8.3 fold decrease between the neutralizing titers of mRNA-1273.214 against BA.4/5 and the ancestral SARS-CoV-2 at day 29, and a 11.5 fold decrease for mRNA-1273.^20^ Using the same method by Khoury et al., 2021^21^ and Hogan et al., 2021,^19^ the initial infection VE against the ancestral strain for mRNA-1273.214 was estimated to be 95.24%. This was driven by the higher GMT level produced against the ancestral strain by mRNA-1273.214 compared to mRNA-1273 (6,422 and 5,287, respectively). VE for the candidate mRNA-1273.222 BA.4/5-adapted bivalent vaccine was approximated using the GMTs of mRNA-1273.214 against BA.1, using the assumption that a BA.4/5-adapted bivalent vaccine would perform similarly against BA.4/5 as a BA.1 vaccine would perform against BA.1 (i.e., 2.6 fold decreased compared to ancestral GMT).^22^ The initial infection VE against the ancestral strain of 95.24% was also assumed for the mRNA-1273.222 vaccine. Waning for all three vaccine boosters was assumed to be the same as waning for primary series vaccination against BA.1/2 (7.3% for infection and 3.4% for severe disease). In order to adjust the VE of the primary series, booster 1, and booster 2 for the Omicron BA.4/5 period, we calculated that VE for the mRNA-1273 vaccine against infection drops to 72% of BA.1 effectiveness using the methods by Hogan et al. 2021,^19^ while VE against severe disease drops to 88% of BA.1 effectiveness.

### Natural Immunity

A review of the data during the pre-Omicron period concluded that the rate of waning of natural immunity is uncertain but likely equal to, or less than, that for vaccine-mediated immunity,^23^ and that the impact of Omicron on the duration of protection from natural immunity is also unclear. For our base case calibration, we assumed conservatively that this waning rate was equivalent to that of vaccine-mediated immunity in the pre-Omicron and Omicron BA.1 periods. We then held the rate constant during the BA.4/5 Omicron period to conduct projections.

### Vaccine Coverage

In this analysis, we attempt to determine the benefit of administering a new bivalent booster to individuals in the US who have received at least a primary series with a currently licensed vaccine. While not yet authorized by the FDA nor recommended by the Centers for Disease Control and Prevention (CDC), we assume this includes receipt of a third booster dose for those who have already received a primary series plus two booster doses, a second booster dose for those who have received only one booster dose, or a first booster dose for those who have received primary series only. The age-specific uptake of the primary series, booster 1, and booster 2 was based on data from the CDC^24^ with final coverage shown in Table 3. For the fall boosters, we used the booster 1 uptake pattern and assumed that individuals are equally likely to receive the booster, regardless of their vaccination history. The age-specific uptake of the fall booster is shown in

**Table 2.** Projected vaccine effectiveness against Omicron BA.4/5 for fall boosters.

| Vaccine | Infection |  | Severe |  |
| --- | --- | --- | --- | --- |
|  | Initial Vaccine Effectiveness | Waning: Monthly Decline | Initial Vaccine Effectiveness | Waning: Monthly Decline |
| **mRNA-1273** | 57.75% | 7.3% | 83.29% | 3.4% |
| **mRNA-1273.214** | 69.55% | 7.3% | 89.73% | 3.4% |
| **mRNA-1273.222** | 87.89% | 7.3% | 96.94% | 3.4% |

**Table 3.** Vaccine uptake by age group: primary series (May 31, 2022); booster 1 (May 31, 2022); and booster 2 (June 15, 2022)

| Age Group (Years) | Primary Series (%) | Booster 1 (%)* | Booster 2 (%)** |
| --- | --- | --- | --- |
| 0-9 | 14.8 | 0 | 0 |
| 10-19 | 54.4 | 21.9 | 0 |
| 20-29 | 65.6 | 34.4 | 0 |
| 30-39 | 67.2 | 36.6 | 0 |
| 40-49 | 74.7 | 44.6 | 0 |
| 50-59 | 81.6 | 54.1 | 17.1 |
| 60-69 | 87.6 | 61.1 | 24.3 |
| 70-79 | 90.7 | 70.1 | 31.4 |
| 80+ | 87.9 | 72.1 | 31.4 |

\* Booster 1 series numbers represent the proportion of the age group that received a booster as a percentage of those that had completed their primary series.
\*\* Booster 2 series numbers represent the proportion of the age group that received a booster as a percentage of those that had completed Booster 1.

Figure 2. For the November mRNA-1273.222 booster, the same uptake pattern was assumed except that vaccination started on November 1, 2022.

**Figure 2.**
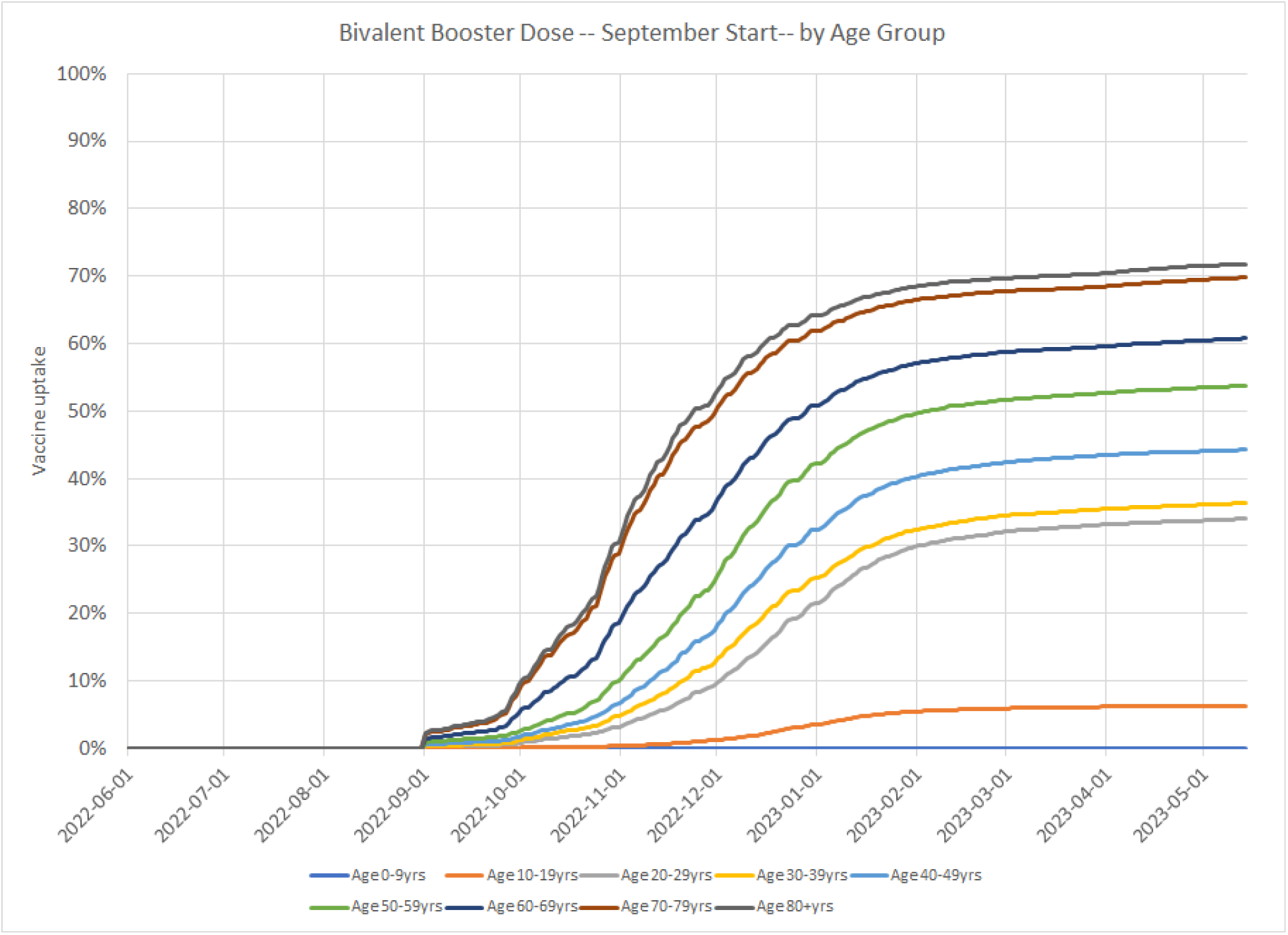
Vaccine uptake for a hypothetical fall booster strategy starting September 1, 2022.

### Consequences of Infections

All patients who develop a symptomatic infection are assumed to move into an infection consequences decision tree (Figure 3). The VE against severe infection is higher than protection against infection alone and this is modeled in the decision tree as a further reduction in hospitalization rates among those who are vaccinated. In sensitivity analysis, the impact of Paxlovid™ (Pfizer; nirmatrelvir/ritonavir) on the risk of hospitalization and death is also taken into account for a percentage of patients in each age stratum who are eligible and ultimately receive the treatment. The key model inputs are displayed in Table 4, with further details provided in the technical appendix.

**Figure 3.**
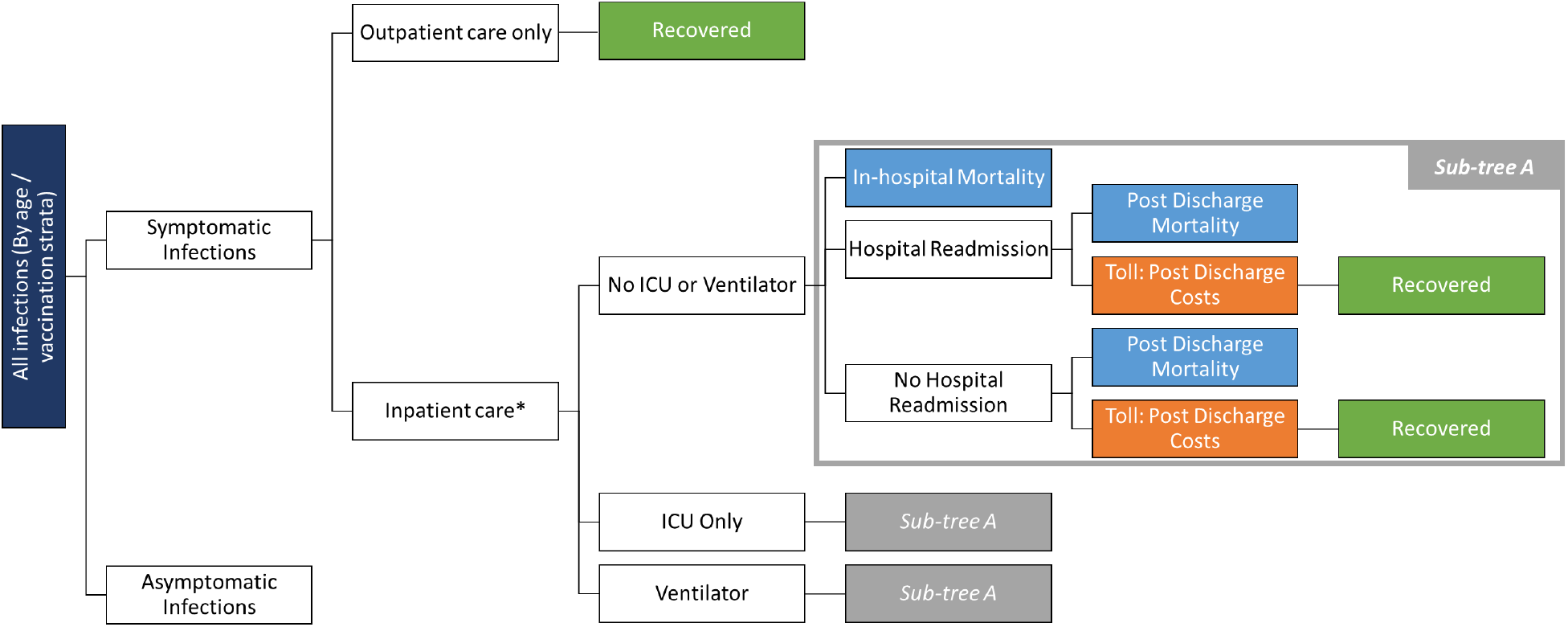
Infection Consequences Decision Tree. *Risk is dependent on eligibility for and receipt of Paxlovid

**Table 4.** Key consequences decision tree model inputs for the base case and sensitivity analyses.

| Model Parameter | Base Case Assumption | Sensitivity Analysis Data/Selections |
| --- | --- | --- |
| Monthly waning rate of mRNA-1273.214 | Same as primary series against Omicron (7.3% for infection, 3.4% for hospitalization) | Decrease of 25%, 50%, and 75% (assumptions) |
| Percentage of Infections with Symptoms | 18-49: 85.3%<br>50-64: 85.1%<br>≥65: 80.8%<br>Reese et al., 2021<sup>25</sup> | ≤18: 39.8%<br>18-39: 50.5%<br>40-49: 67.5%<br>50-64: 66.8%<br>≥65: 66.2%<br>Ma et al., 2021<sup>26</sup> |
| Age-specific hospitalization rates in the unvaccinated population | 18-49: 1.25%<br>50-64: 3.45%<br>≥65: 15.71%<br>Reese et al., 2021<sup>25</sup>,<br>adjusted by Wang et al., 2022<sup>27</sup> | 18-49: 2.6%<br>50-64: 7.2%<br>≥65: 22.4%<br>Reese et al., 2021<sup>25</sup> |
| Paxlovid™ treatment | 0% | Eligible: 15.1%<br>Of those eligible, utilization rate:<br>18-64 years old: 17.2%<br>≥ 65 years old: 2.1%<br>Qasmieh et al., 2022<sup>28</sup> |
| Masking | 0% at end of July 2022 | 9%<sup>11</sup> at end of July 2022 and remains at this level for an 8-month period, then reduced to 0%. |

### Analyses Conducted

We conducted a base case analysis to compare the clinical impact of the three potential strategies for fall 2022 to no additional boosters: 1) licensed mRNA-1273 (ancestral monovalent) booster starting in September; 2) candidate mRNA-1273.214 (ancestral + BA.1) booster starting in September; and 3) candidate mRNA-1273.222 (ancestral + BA.4/5) booster starting in November.

We then recalibrated the model and varied the assumptions about waning of natural immunity and vaccine-mediated immunity to determine how waning would impact projections of infections for the fall. We altered the transmissibility of the virus to recalibrate to the IHME infection counts. From June onwards, however, we did not change the transmissibility parameter input from the base case. We tested the impact of: 1) reducing the waning of natural immunity by 50%, implying that it wanes at half the rate of vaccine-mediated immunity; 2) increasing the waning rate of vaccine immunity to 110% of the base values; and 3) decreasing the waning rate of vaccine immunity by 90% of the base values. We show results for the mRNA-1273.214 booster only. Using our original calibration, we also increased and decreased the virus transmissibility during the projection period by 10% and delayed the increase in transmissibility by one month. We decreased the uptake of the fall booster by 25% across all age groups. We present the impact of these changes on projected infections.

The base case assumed that the VE monthly waning rate against BA.4/5 for the mRNA-1273.214 booster was the same as the weighted primary series waning rate calculated for BNT162b2, mRNA-1273, and AD26.CO2.S. However, there is emerging evidence suggesting that due to the bivalent composition of mRNA-1273.214, the duration of protection is extended (i.e., waning is reduced).^29^ To determine the impact of decreased waning due to bivalent vaccines, three sensitivity analyses were conducted on the waning rate of mRNA-1273.214: a 25%, 50%, and 75% decrease in waning compared to the primary series.

Finally, we conducted a range of sensitivity analyses by varying our decision tree inputs as shown in Table 4.

## 3. RESULTS

The average VE, by vaccination status, as projected by the model in the base case for the bivalent mRNA-1273.214, is shown in Figure 4 for those 80 years and above as an example. In the graph, solid lines represent VE against infection and dotted lines represent VE against severe disease. By September 2022, the model predicts that those who received only the primary series or booster 1 have no vaccine-mediated immunity against infection, while booster 2 VE is below 30%. VE against severe diseases lasts longer, but it is projected to be below 50% for those receiving the primary series only. Among those who receive a mRNA-1273.214 booster, protection against infection and severe infection is boosted to 70% and 90%, respectively.

**Figure 4.**
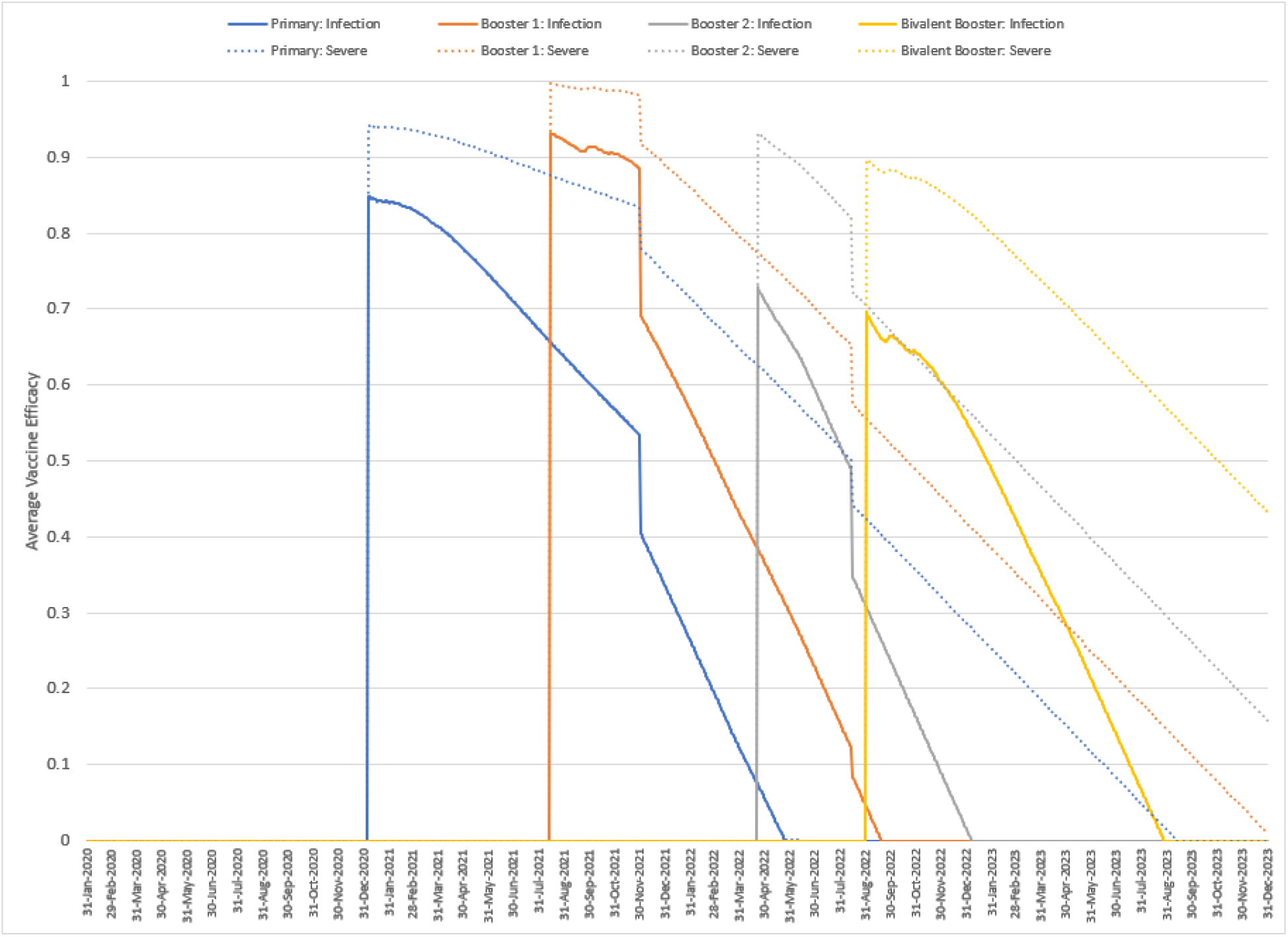
Model projected vaccine efficacy against infection and severe disease for primary series, booster 1, booster 2, and fall booster (mRNA-1273.214) groups.

The model-projected numbers of infections with and without a fall booster are shown in Figure 5. From September 2022 to February 2023 where healthcare systems are the most pressured, mRNA-1273 is predicted to prevent 34% of infections compared to no booster, mRNA-1273.214 prevents 40%, while mRNA-1273.222 prevents only 18% due to the delay in starting to boost. Given that the model predicts no protection against infection for many individuals, the administration of any fall booster reduces future infections.

**Figure 5.**
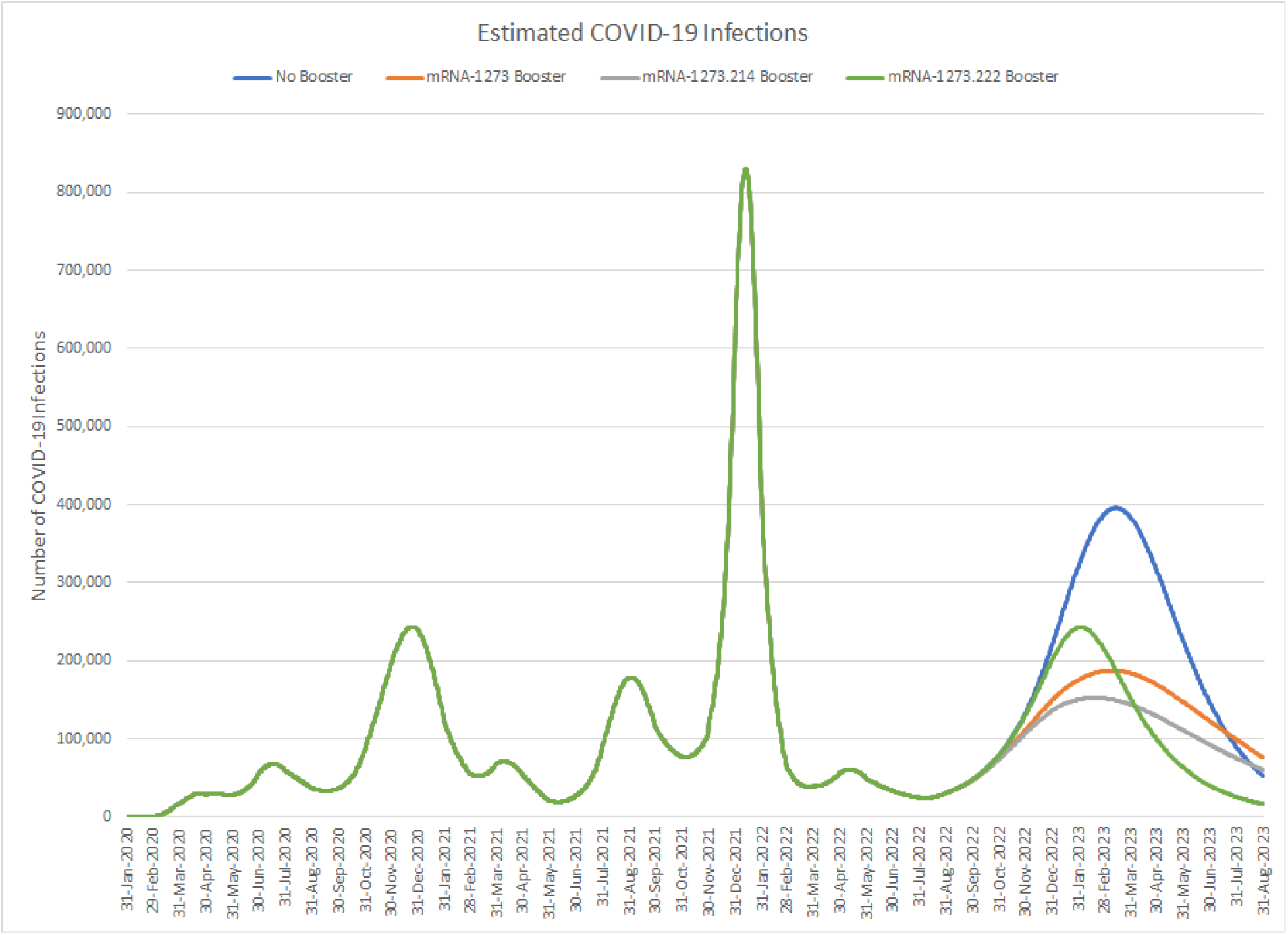
Base case results: Estimated COVID-19 Infections with and without a fall booster strategy (mRNA-1273 or mRNA-1274.214 starting in September; mRNA-1273.222 starting in November)

Table 5 displays the cumulative number of infections predicted for each scenario by month and the percent change for each booster compared to no booster. We have assumed that BA.4/5 will persist for the entire 6-month time horizon, however, if a new variant becomes dominant within the time period the relative effectiveness of the vaccines may change. If we further assume that BA.4/5 will persist over one year, the licensed mRNA-1273 booster is projected to reduce infections by 36%, the candidate bivalent mRNA-1273.214 by 47%, and the candidate mRNA-1273.222 by 45% compared to no boosters given in the fall.

**Table 5.** The cumulative number of infections predicted by the model by month for all booster scenarios.

|  | Month 1<br>Sept 2022 | Month 2<br>Oct 2022 | Month 3<br>Nov 2022 | Month 4<br>Dec 2022 | Month 5<br>Jan 2023 | Month 6<br>Feb 2023 | Month 12<br>Aug 2023 |
| --- | --- | --- | --- | --- | --- | --- | --- |
| **Cumulative number of infections** |  |  |  |  |  |  |  |
| **No**<br>**Booster** | 1,161,443 | 3,112,392 | 6,271,954 | 11,722,566 | 20,262,504 | 30,373,482 | 72,693,116 |
| **mRNA**<br>**1273** | 1,154,092 | 3,039,360 | 5,845,524 | 9,926,195 | 15,040,778 | 20,160,457 | 46,374,970 |
| **mRNA**<br>**1273.214** | 1,152,428 | 3,023,085 | 5,753,905 | 9,566,330 | 14,072,523 | 18,336,591 | 38,500,425 |
| **mRNA**<br>**1273.222** | 1,161,443 | 3,112,392 | 6,240,011 | 11,404,660 | 18,480,882 | 25,019,073 | 40,069,698 |
| **Percent change from no booster** |  |  |  |  |  |  |  |
| **mRNA**<br>**.1273** | -1% | -2% | -7% | -15% | -26% | -34% | -36% |
| **mRNA**<br>**1273.214** | -1% | -3% | -8% | -18% | -31% | -40% | -47% |
| **mRNA**<br>**1273.222** | 0% | 0% | -1% | -3% | -9% | -18% | -45% |

As with infections, starting to boost earlier (i.e., September) prevents more hospitalizations over the 6-month period than starting to boost two months later with a more effective vaccine: 58%, 66%, and 46% for mRNA-1273, mRNA-1273.214, and mRNA-1273.222, respectively. The impact of different boosters on the expected number of hospitalizations is shown in Figure 6. Overall, using a booster prevents hospitalizations mainly because it prevents infections in people in all strata, including those who do not receive the booster through the effect of herd immunity. There is some additional benefit from increasing protection against severe disease in those who received the fall booster as well. Assuming BA.4/5 will persist over one year, the licensed mRNA-1273 booster is projected to reduce hospitalizations by 47%, the candidate bivalent mRNA-1273.214 by 57%, and the candidate mRNA-1273.222 by 55% compared to no boosters given in the fall.

**Figure 6.**
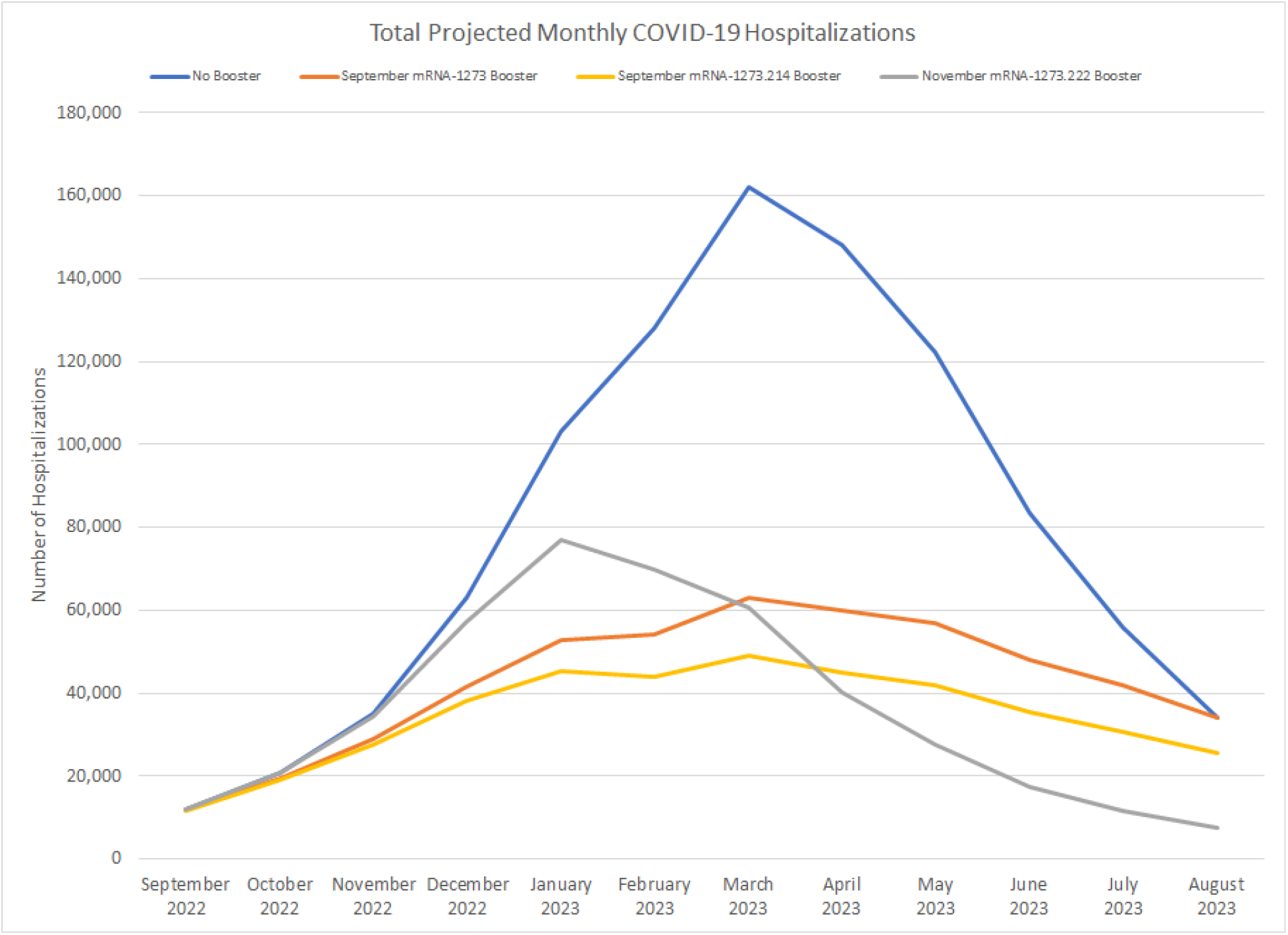
Estimated COVID-19 hospitalizations with and without a fall booster strategy (mRNA-1273 or mRNA-1274.214 starting in September; mRNA-1273.222 starting in November)

Our sensitivity analyses with the SEIR model demonstrate that assumptions on the waning of natural immunity, transmissibility and vaccine coverage impact the projected number of infections and the effectiveness of a fall booster as shown in Figure 7, Figure 8 and Figure 9. When we varied the rate of natural immunity and the vaccine-induced immunity waning rates during our calibration period, natural immunity waning had the largest impact on the projected number of cases this fall (Figure 7). While the number of cases projected decreases with stronger natural immunity, the mRNA-1273.214 is still effective when assuming half the waning rate of natural immunity and reduces the number of infections by 36% compared to no fall booster. However, even with this higher level of natural immunity, we can still achieve the same magnitude of number of infections starting September 2022 as predicted in the base case by increasing the transmissibility of the virus by 11%.

**Figure 7.**
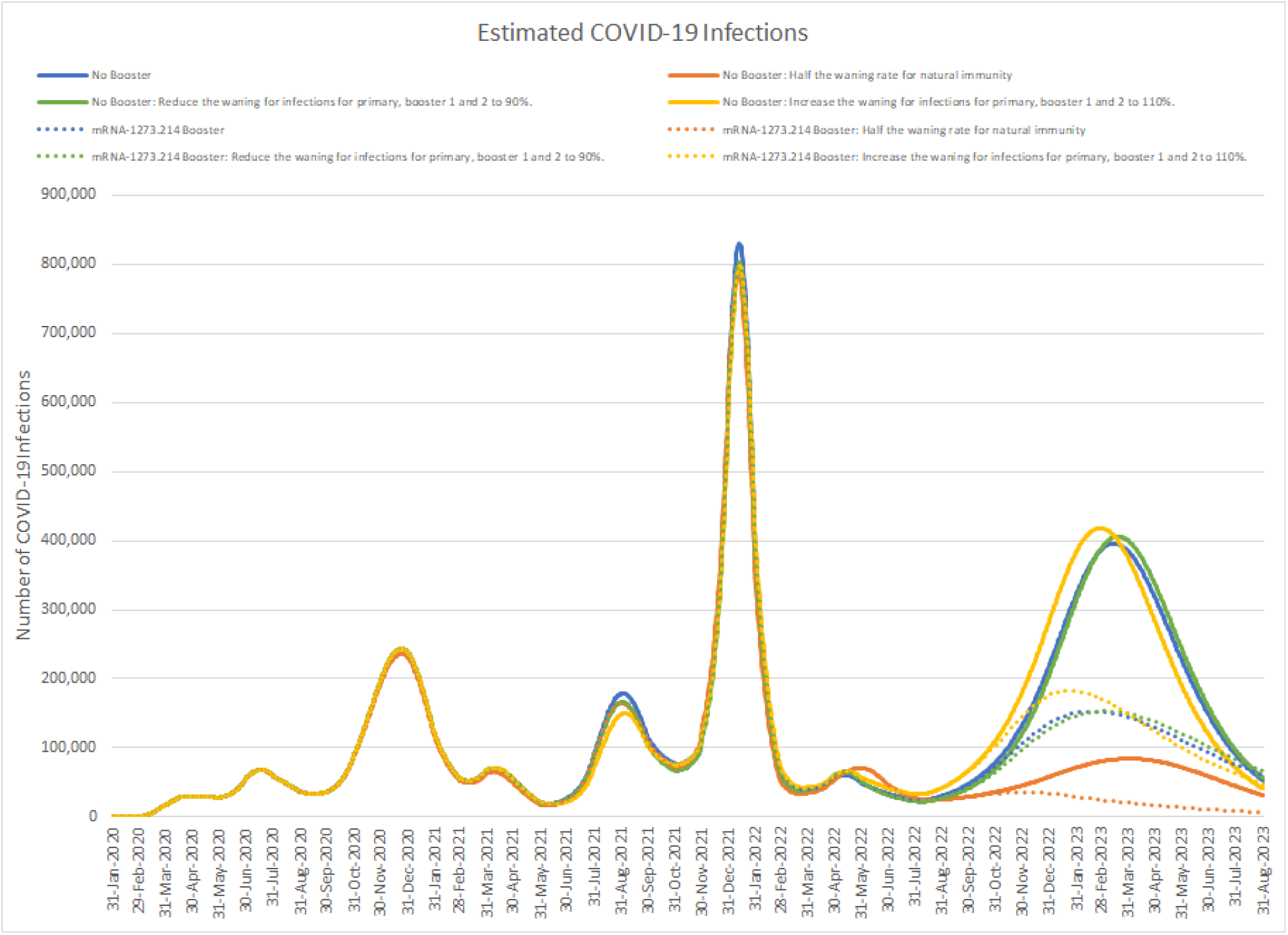
Impact of changing calibration assumptions on the estimated number of infections projected with no fall booster strategy and the mRNA-1273.214 September strategy.

**Figure 8.**
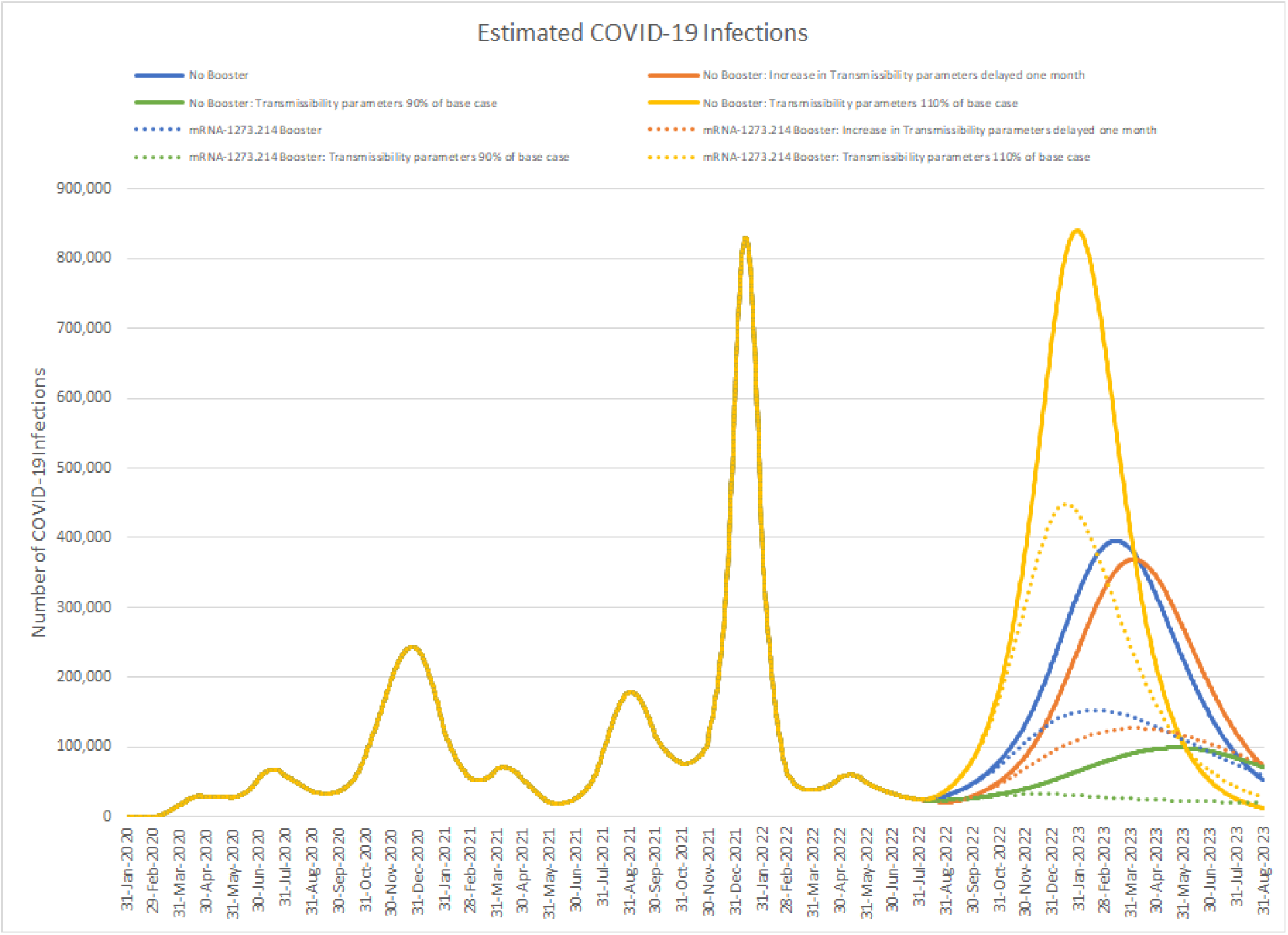
Impact of changing the transmissibility of the virus or delaying the increase in transmissibility caused by a new variant by 1 month.

**Figure 9.**
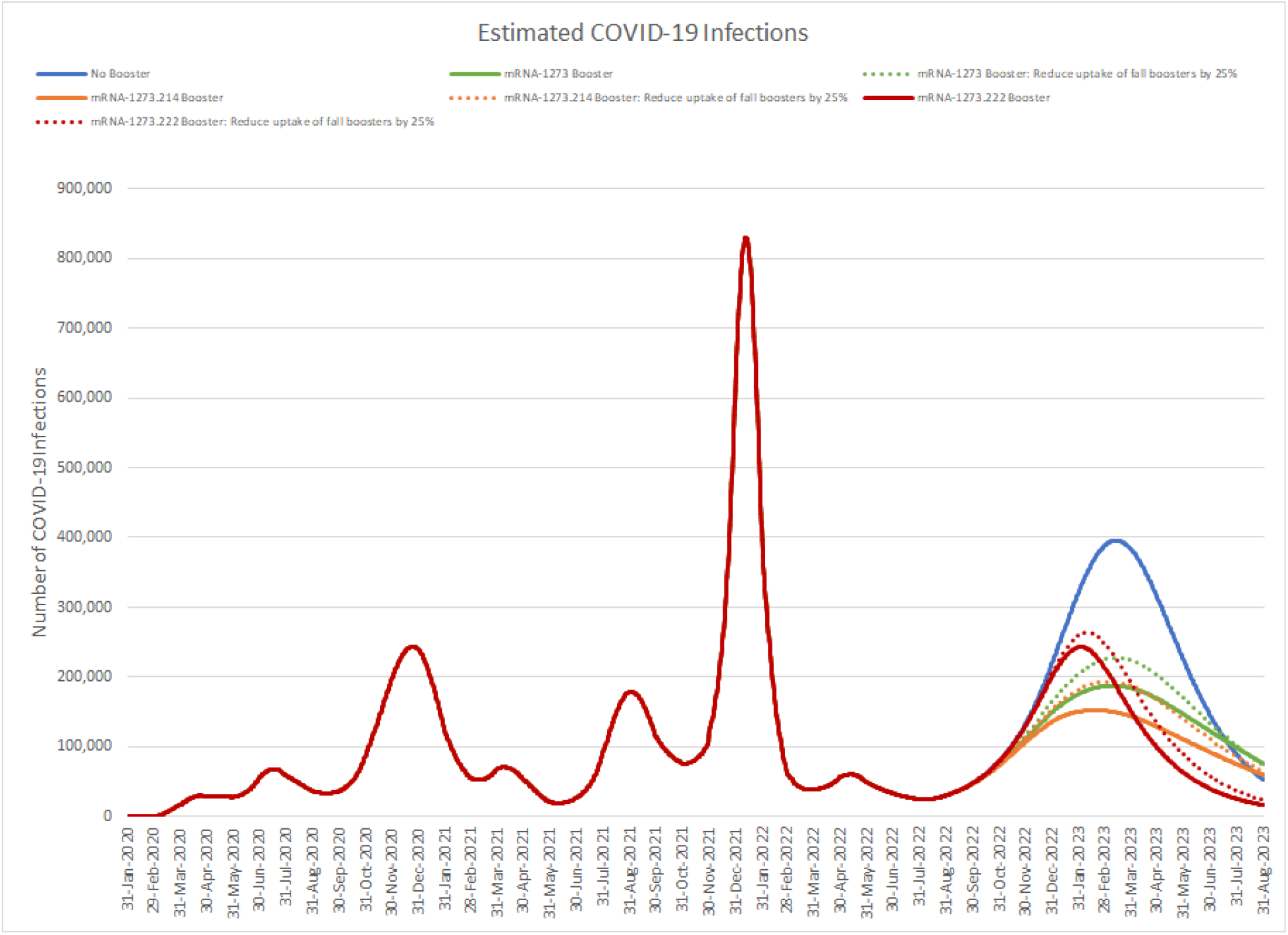
Impact of reducing the coverage of a fall booster strategy to 25% of base case on the number of infections.

When we used our base case calibration but increased or decreased the transmissibility of the future VOC, the absolute projected number of infections with no booster increased or decreased accordingly (Figure 8). The percent decreases in 6 months of cases when using the mRNA-1273.214 booster compared to no booster are 33% and 36%, respectively, which compares to 40% in the base case. If the increase in transmissibility of the virus is delayed from August 15 to September 15, 2022, but the fall boosters are still administered beginning in September, the decrease in number of infections with mRNA-1273.214 compared to no booster is 42%. If uptake with the fall booster is 25% of what is predicted in base case, the decrease in the number of infections is lower: 27% with mRNA-1273, 32% with mRNA-1273.214 and 14% with mRNA-1273.222 (Figure **9**). Decreasing the waning rate (i.e., increasing the duration of protection) of the mRNA-1273.214 booster by 25%, 50%, and 75% of the base case values prevented 41%, 42%, and 43% of infections, respectively, compared to no booster. Likewise, hospitalizations prevented were 68%, 69%, and 71%, respectively. Although increasing masking to 9% from 0% decreased the absolute number of infections and hospitalization both by 41%, it had minimal impact on the proportion prevented compared to no booster: the percentage decrease remained at 34%, 40%, and 18% for infections, and 43%, 48%, and 25% for hospitalizations, for mRNA-1273, mRNA-1273.214, and mRNA-1273.222, respectively.

Using an alternate source to inform the proportion of infections that are symptomatic or the age-specific hospitalization rates had no effect on total number of infections prevented by each of the boosters; likewise, including Paxlovid™ treatment only affected number of hospitalizations. Percentage decrease compared to no booster for each vaccine are presented in Table 6.

**Table 6.**
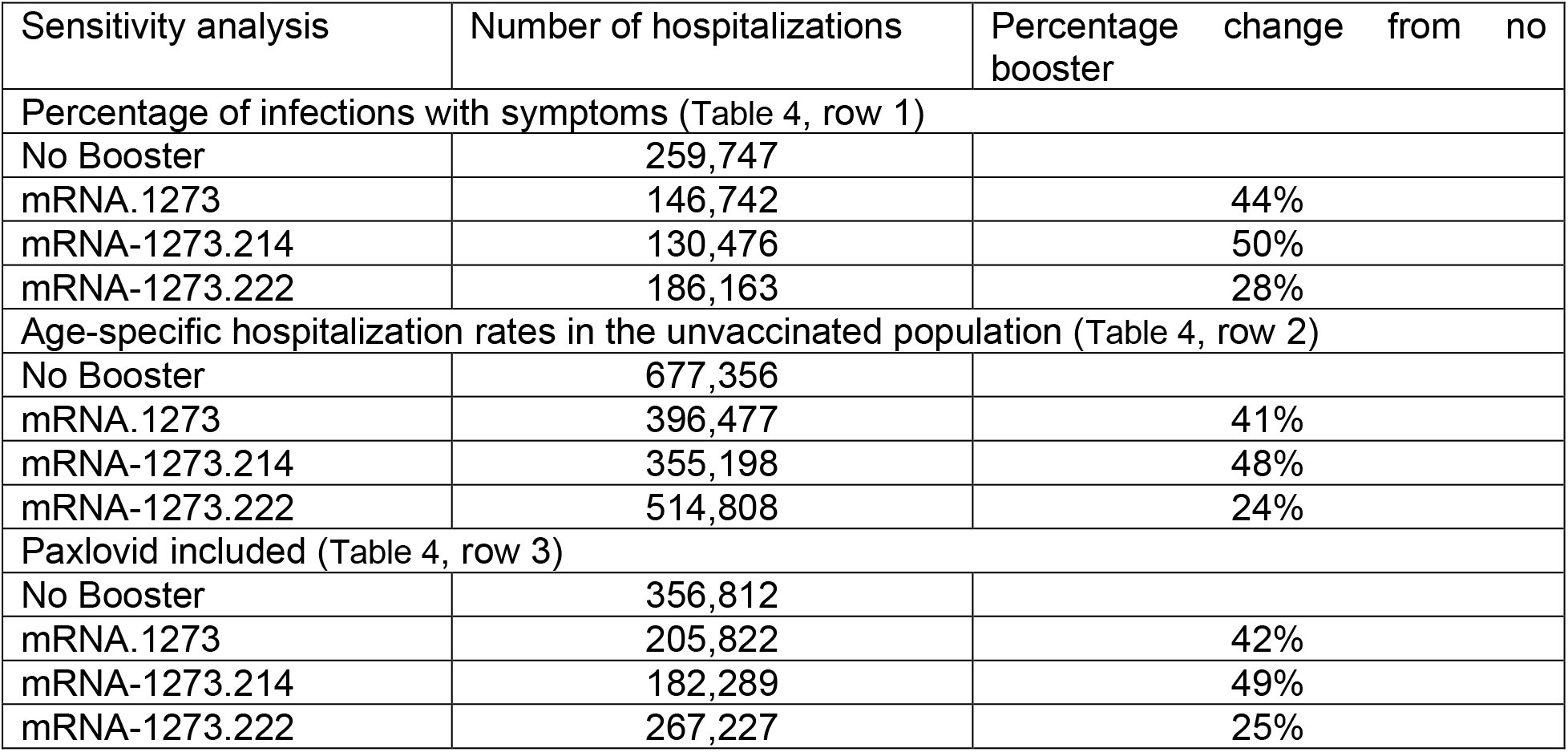
Sensitivity analyses: Hospitalization results over 6 months.

| Sensitivity analysis | Number of hospitalizations | Percentage change from no booster |
| --- | --- | --- |
| Percentage of infections with symptoms (Table 4, row 1) |  |  |
| No Booster | 259,747 |  |
| mRNA.1273 | 146,742 | 44% |
| mRNA-1273.214 | 130,476 | 50% |
| mRNA-1273.222 | 186,163 | 28% |
| Age-specific hospitalization rates in the unvaccinated population (Table 4, row 2) |  |  |
| No Booster | 677,356 |  |
| mRNA.1273 | 396,477 | 41% |
| mRNA-1273.214 | 355,198 | 48% |
| mRNA-1273.222 | 514,808 | 24% |
| Paxlovid included (Table 4, row 3) |  |  |
| No Booster | 356,812 |  |
| mRNA.1273 | 205,822 | 42% |
| mRNA-1273.214 | 182,289 | 49% |
| mRNA-1273.222 | 267,227 | 25% |

## 4. DISCUSSION

We conducted an analysis using mathematical modeling to compare three different booster strategies that involved providing an additional Moderna booster dose in Fall 2022 to adults who had previously been vaccinated with at least a primary series. The options were: (1) the licensed monovalent mRNA-1273; (2) the candidate bivalent mRNA-1273.214; or (3) the candidate bivalent mRNA-1273.222 updated for BA.4/5. We compared all of these to the counter-factual of no additional boosting. As additional development time would be required for the mRNA-1273.222 bivalent booster, we assumed that boosting would start 2 months later than boosting with the other two vaccines.

Our analysis demonstrated that vaccinating with the bivalent mRNA-1273.214 was more effective over a 6-month period in preventing infections with a BA.4/5 subvariant than the tailored vaccine, simply because it could be deployed earlier. By September 2022, our model predicts that vaccine-induced protection against infection is quite low, especially in those who have only received a primary series or one booster. The earlier deployment allows reduction in circulating virus in the population sooner and prevents an exponential rise in the number of infections. This result was consistent with one of our sensitivity analyses in which the mRNA-1273.214 booster was even more effective in reducing infections if the BA.4/5 wave is delayed by 1 month. Focusing on the initial 6-month period when healthcare systems are the most pressured, vaccinating with either the monovalent mRNA-1273 or the bivalent-1273.214 is more effective than delaying to boost with a BA.4/5-specific vaccine (mRNA-1273.222). Reduction in infections is important because they decrease the number of hospitalizations across the population, even amongst those who are not vaccinated or up to date on the recommended boosters.

Our analysis also demonstrates that the candidate bivalent mRNA-1273.214 has substantial incremental benefits in terms of infections and hospitalizations compared to the licensed monovalent mRNA-1273. Based on antibody titers, we estimated a 11.8% difference in initial VE against infection, and 6.4% difference against severe disease against BA.4/5, using the same waning rate over time. Although it is not known how these vaccines will protect against future VOCs, current immunogenicity data suggests that bivalent vaccines offer a broader protection across current VOCs and are potentially more durable, compared to first generation monovalent vaccines. Using neutralizing antibody titer levels, mRNA-1273.214 was shown to elicit higher GMTs at day 29 compared to mRNA-1273 against ancestral and all variants tested (Alpha, Beta, Delta, Gamma, BA.1, and BA.4/5).^20^

Data on VE for mRNA-1273, BNT162b2, and AD26.COV2.S administered prior to June 15, 2022 were obtained from published meta-analyses, adding strength to our data. However, there are a lack of VE data for BA.4/5 since a new VOC is often dominating by the time the study data are disseminated. Extensive work has been done on algorithms to predict, with some degree of certainty, VE against current strains based on GMT levels.^18^ The VEs against BA.4/5 for the fall booster mRNA-1273, candidate mRNA-1273.214 and mRNA-1273.222 bivalent vaccines are not available, and relative GMT levels were used instead. Therefore, all fall booster VEs were derived using the same method, which is another strength of our analysis. Waning rates for the bivalent vaccines were assumed to be the same as monovalent primary series rates, which is a conservative estimate, as there is emerging evidence suggesting a bivalent vaccine maintains the duration of protection longer.^29^ To test the impact of this assumption, a series of sensitivity analyses were performed looking at the difference in infections and hospitalization between mRNA-1273 and mRNA-1273.214 if the bivalent waning rate was decreased. Sensitivity analyses decreasing the waning rate of mRNA-1273.214 by 25-75% (i.e., assuming more durable protection) of the base case value increased the number of prevented infections and hospitalizations compared to no booster from 40% to 41-43% and from 57% to 68%-71%, respectively. Therefore, our base case benefits of mRNA-1273.214 over mRNA-1273 may be an underestimate of actual numbers of infections and severe disease prevented. Varying the estimates for proportion of infections that are symptomatic, age-specific hospitalization rates, or including Paxlovid had minimal impact on the percentage decreases in hospitalization, suggesting they were not key drivers of the model results. Increasing the proportion masking decreased cases of infection and hospitalizations compared to the base case, but did not change the percentage reduction in infections and hospitalizations compared to no booster with masking.

With the emergence of new variants, key characteristics of the virus that affect estimates of spread and clinical impact, such as transmissibility, disease severity, risk of reinfection, and vaccine effectiveness also evolve, making estimation of these parameters difficult, especially in heterogeneous populations. While mathematical models have had some success at predicting the course of infections across a 1-month time horizon, longer-term predictions have been challenging.^12^ Longer-term projections also necessitate assumptions about the emergence of future VOCs and their characteristics, as well as the population’s behavior to protect themselves in reaction to their emergence, which are inherently difficult to predict over the long-term. As we demonstrated in our sensitivity analyses, higher proportions of natural immunity in the population will blunt the number of infections caused by future variants. However, it has been observed that natural immunity obtained from past variants such as delta, may be significantly reduced with newer variants such as Omicron BA.1.^30^ It is not possible to know how well infections with past variants protect against future variants. Even with different levels of natural immunity, the magnitude of infection incidence also depends on the transmissibility of future VOCs. Given the unknowns, we acknowledge that our projections will not represent the incidence that will be seen in a heterogeneous and geographically diverse country like the US. However, our projections of the impact of the vaccine have shown that a fall booster is useful under multiple scenarios to prevent both illness and hospitalizations.

Future work will include updating the model and estimates with data as they emerge, conducting a cost-effectiveness analysis on the use of different boosters, as well as examining the impact of adverse events such as vaccine-associated compared to COVID-19-associated myocarditis.

## 5. CONCLUSION

We conclude that providing a fall booster prevents infections and hospitalizations compared to no further vaccination. Furthermore, vaccinating with the next generation bivalent vaccine mRNA-1273.214 is predicted to reduce infections by 40% compared to no booster over a 6-month period, while the monovalent mRNA-1273 will only reduce them by 34%. Compared to boosting with mRNA-1273.214 in September 2022, delaying boosting by 2 months until a BA.4/5 vaccine (mRNA-1273.222) is available results in only 18% of infections being prevented across the 6-month period. Therefore, we conclude that there is no advantage to delay boosting until a BA.4/5 vaccine is available; earlier boosting with mRNA-1273.214 will prevent the most infections and the most hospitalizations.

## Supporting information

Technical Appendix

## Data Availability

All data produced in the present work are contained in the manuscript and supplemental materials.

## Author Contributions

AL, KF, MK, MM, PB, NV and MW were involved in study design and interpretation of the analysis. MM programmed the model with quality assurance by MK, AL, and KF. All authors were involved in model estimation. MM, MK and KF conducted the analysis. AL, KF and MK wrote the initial draft of the manuscript, and all remaining co-authors critically revised the manuscript and approved the final version.

