## Supplementary material for "The Potential Clinical Impact of Implementing Different COVID-19 Boosters in Fall 2022 in the United States": Technical Appendix

1. Technical Appendix

### 1.1. Dynamic Model Structure

The basic structure of the dynamic transmission model is presented in Figure 1 in the main text, while the model equations are shown in section in this appendix.

All individuals in the simulation begin in the susceptible compartment in the unvaccinated stratum except for a few individuals representing imported COVID-19 cases (over the first 30 days) that are required to start the pandemic. Individuals move between the susceptible and exposed compartments according to the force of infection (see equations in Section 1.4).

During the simulation, individuals move between the model strata (rows in Figure 1) according to uptake rates for the vaccine (primary series; booster 1; booster 2; fall booster). The fall booster stratum is used for projections that explore the impact of boosting with either mRNA-1273, the candidate vaccine mRNA-1273.214, or the candidate vaccine against BA.4/5, mRNA-1273.222.

Once exposed to an infection, an individual develops a latent infection (exposed compartment / pre-infectious state), which cannot be transmitted. As the infection progresses, individuals lose latency and move to the infectious state, which can manifest as an asymptomatic infection or one with clinical symptoms. The rate of loss of latency is a function of the average latent period. Similarly, the loss of infectiousness is a function of the average duration of a COVID infection. The incidence of infection is measured as the movement into the infectious state compartment. Following recovery from an infection, individuals move into a recovered compartment in which they have natural immunity and cannot develop another infection. If the individuals are in one of the vaccinated strata, being in the recovered health state corresponds to having had a breakthrough infection with hybrid immunity. In the recovered state, individuals’ natural immunity wanes until they return to the susceptible state. The rate of waning of natural immunity is the same regardless of vaccination status.

The time step of the simulation was set to be 1 day.

#### 1.1.1. Assumptions

The following key assumptions were made in the design of the dynamic model:

1. Contacts between individuals in different age groups were based upon a specified contact matrix. Otherwise, contact between individuals was assumed to be random.
2. An initial 3,136 infections during the first 30 days of the simulation were assumed to seed or start the pandemic. This number is equal to the number of cases estimated by the Institute for Health Metrics and Evaluation (IHME) over the first 30 days.^11^ The infections were equally spread out over the first 30 days of the simulation and across the age groups (Table 1).
3. Immunocompromised individuals are not explicitly modelled in this analysis.
4. Individuals under 18 years of age are modelled, however, their vaccination status does not change after May 30, 2022 and they are not eligible to receive a fall booster as this analysis focuses on adults only.

Table 1. Number of infections (over 30 days) used to seed the simulation by age group

| **Age Group (Years)** | **Infections (over 30 days)** |
| --- | --- |
| 0-9 | 348 |
| 10-19 | 348 |
| 20-29 | 349 |
| 30-39 | 349 |
| 40-49 | 349 |
| 50-59 | 349 |
| 60-69 | 348 |
| 70-79 | 348 |
| 80+ | 348 |
| **Total** | **3,136** |

### 1.2. Dynamic Model Inputs: January 2020 to May 2022

The purpose of simulating the pandemic from January 2020 to May 2022 was to allow the model to estimate a number of parameters for the end of May 2022. These include the number of people in the R (recovered) compartments, the proportions of the population that are in each of the model vaccine strata, and the average vaccine effectiveness in each vaccine/booster stratum, given staggered injections across the days and waning of vaccine protection.

#### 1.2.1. Vaccine Coverage

A proportion of the population moves into the primary series and booster model strata according to vaccine uptake data from the CDC, as illustrated in Figure 1 to Figure 3.^24^

Figure 1. Percent of the population who have completed their primary series, by age group

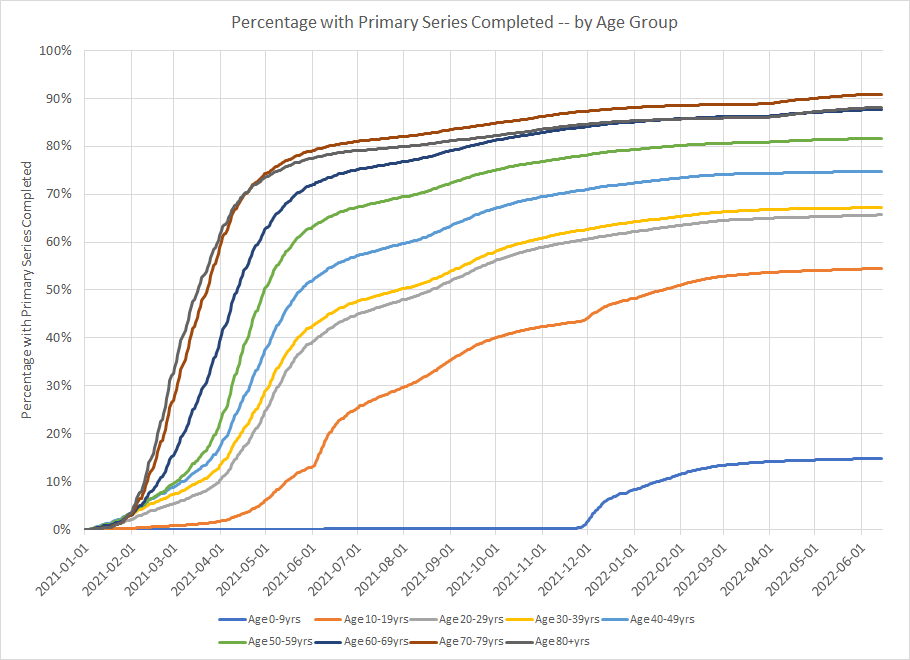

Figure 2. Percent of the population who received their primary series that also received a booster, by age group

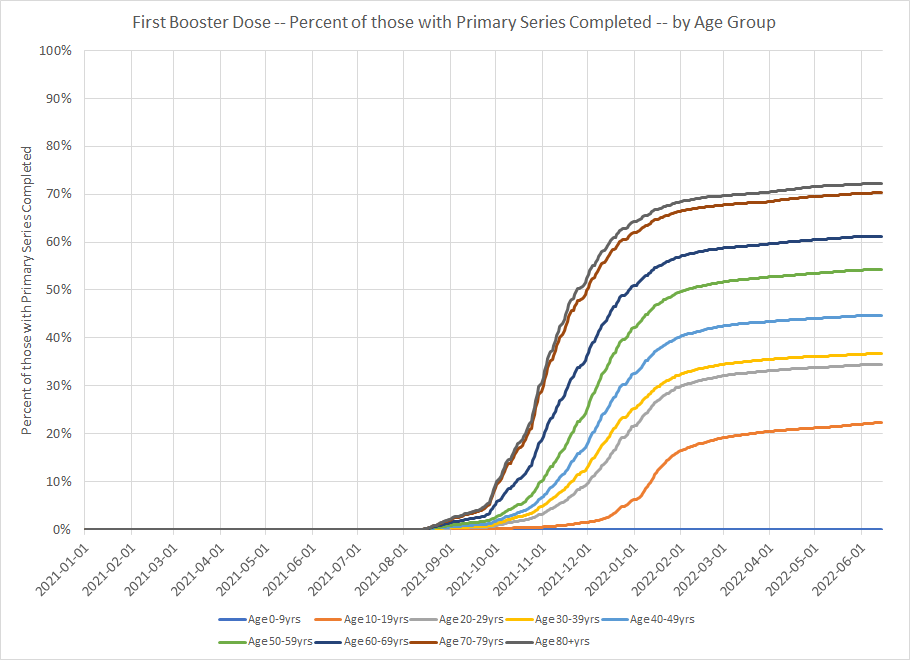

Figure 3. Percent of the population who received two boosters, by age group

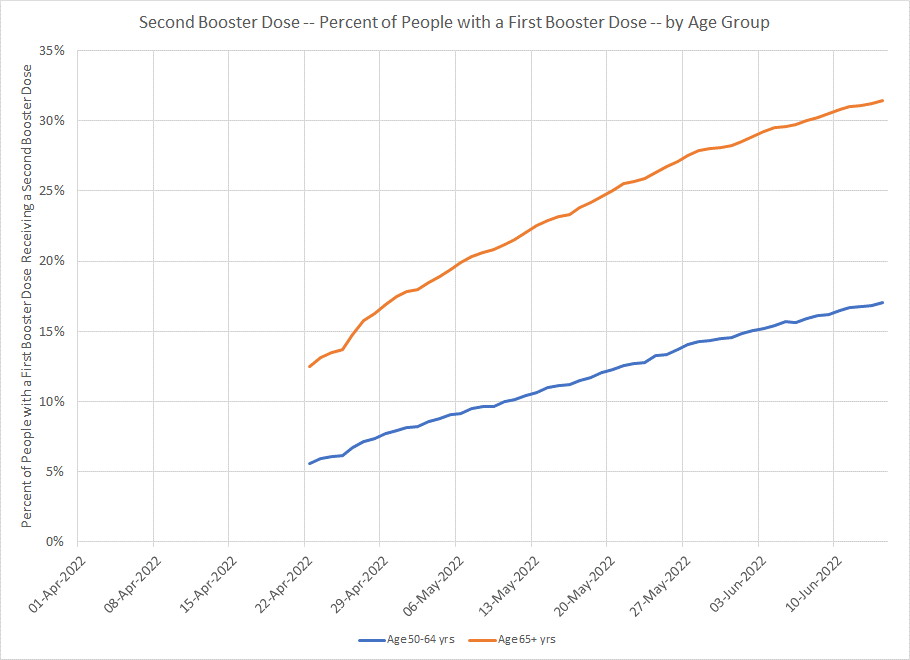

#### 1.2.2. Latent and Infectious Periods

The length of time spent by infected individuals exposed to the virus before they become infectious was assumed to be 3 days.^31^

The length of time spent by an individual in an infectious state was assumed to be 7 days.^31,32^

#### 1.2.3. Natural Immunity

The rate of waning of natural immunity following COVID-19 infection has not been determined and is complicated by confounding factors and the change in circulating variants over time. While it is certain that infection confers immunity, it is unclear how long that immunity lasts. Following a review of the efficacy and duration of natural immunity based on data published prior to January, 2022, Pilz et al.^23^ concluded, “Taken together, observational studies indicate that natural immunity offers equal or greater protection against SARS-C0V-2 infections compared to individuals receiving two doses of a mRNA vaccine, but data are not fully consistent.” Data on the impact of Omicron on natural immunity are now emerging but have not yet been summarized systematically. For the base case, we assumed that the waning rate of natural immunity was equivalent to the waning rate for vaccine-mediated immunity for the pre-Omicron and Omicron (BA.1) period. For the Omicron BA.4/5 period the waning rate was set equal to the BA.1 period. While vaccine-mediated immunity declined when a new VOC period began, we did not alter the proportion of the population who were in the recovered states and therefore have natural immunity.

#### 1.2.4. Force of infection

The force of infection is a function of the number of susceptible people in the population as well as the rate of effective contacts between susceptible and infected individuals. Effective contacts are a function of the rate of contact, which is driven by an age-specific contact matrix, and the transmissibility of the virus per contact. For individuals in the various vaccine strata, the force of infection is reduced based on the average vaccine effectiveness of the cohort.

Model inputs for the force of infection are described in the sections below.

**Population Size**

The size of the US population in 2019 by age was obtained from the United States Census Bureau.^33^

Table 2. The model population size

| **Age Group (Years)** | **Number** |
| --- | --- |
| 0-9 | 39,772,578 |
| 10-19 | 41,852,838 |
| 20-29 | 45,141,956 |
| 30-39 | 44,168,826 |
| 40-49 | 40,319,374 |
| 50-59 | 42,354,542 |
| 60-69 | 38,026,147 |
| 70-79 | 23,681,097 |
| 80+ | 12,922,165 |
| **Total** | 328,239,523 |

**Mixing Patterns / Contact Matrices**

Data on the age-specific mixing patterns in the general population for the United States were obtained from Prem and colleagues.^34^ In the published contact matrices, the population was partitioned into 5-year age bands, and all individuals aged 75 years and older were grouped together. For this analysis, the age-specific mixing patterns were first converted into 10-year age bands. Then, we assumed symmetry between the age groups (i.e., an effective contact between someone in age group *i* with someone from age group *j* is the same as an effective contact between someone in age group *j* with someone from age group *i),* weighted by the population estimates in age groups *i* and *j*.

Thus, $c_{ij}=\frac{1}{N_{j}}\times\frac{\left( c_{ij}^{*}\times N_{j} \right)+\left( c_{ji}^{*}\times N_{i} \right)}{2}$

Where:

$c_{ij}$ is the number of effective contacts between someone in age group *i* with someone from age group *j*

$c_{ij}^{*}$ is the number of effective contacts between someone in age group *i* with someone from age group *j* based on the original age-specific mixing patterns were first converted into 10-year age bands

$N_{i}$ is the population size in age group *i*.

The base contact matrix is presented in Table 3.

Table 3. Base contact matrix used in the model before applying scaling factors

| **Age Group of**  **Participant (Years)** | **Age Group of Contact (Years)** | | | | | | | | |
| --- | --- | --- | --- | --- | --- | --- | --- | --- | --- |
|  | **0-9** | **10-19** | **20-29** | **30-39** | **40-49** | **50-59** | **60-69** | **70-79** | **80+** |
| **0-9** | 5.0604 | 1.2670 | 0.7715 | 1.7630 | 1.0016 | 0.7916 | 0.5682 | 0.4429 | 0.7981 |
| **10-19** | 1.3333 | 11.7181 | 1.6963 | 1.4153 | 2.1288 | 1.3544 | 0.4907 | 0.6714 | 1.2603 |
| **20-29** | 0.8757 | 1.8296 | 5.6961 | 2.5122 | 2.0989 | 1.7911 | 0.6032 | 0.3591 | 0.5408 |
| **30-39** | 1.9578 | 1.4936 | 2.4581 | 4.3860 | 2.8645 | 1.8419 | 0.9122 | 0.5834 | 0.9352 |
| **40-49** | 1.0153 | 2.0508 | 1.8747 | 2.6148 | 3.7184 | 1.9958 | 0.6981 | 0.8008 | 1.2605 |
| **50-59** | 0.8429 | 1.3706 | 1.6805 | 1.7662 | 2.0966 | 2.8073 | 0.8874 | 0.6151 | 1.0491 |
| **60-69** | 0.5433 | 0.4458 | 0.5081 | 0.7853 | 0.6584 | 0.7967 | 1.4255 | 0.7012 | 0.6706 |
| **70-79** | 0.2637 | 0.3799 | 0.1884 | 0.3128 | 0.4703 | 0.3439 | 0.4367 | 0.8033 | 0.8743 |
| **80+** | 0.2593 | 0.3891 | 0.1548 | 0.2736 | 0.4040 | 0.3201 | 0.2279 | 0.4771 | 0.7925 |

Note: Since the population size of the age groups are not of equal size, the transformed contact matrix is not strictly symmetric.

During the pandemic, the rate of contact was reduced by behaviors such as social distancing and mask use. The magnitude of the impact is estimated as described in the next section.

**Social Distancing / Mask Use**

Data on social distancing patterns and mask use were obtained from the IHME and used to adjust the base contact matrix based on these factors.^11^ Monthly estimates on social distancing patterns and mask use were obtained from the IHME for the time period February 2020 through June 2022, inclusive, and are shown in Figure 4 and Figure 5.

Mask use represents the percentage of the population who say they always wear a mask in public. Daily estimates of mask use were obtained from IHME.^11^ It was assumed that 100% mask usage is associated with a 30% reduction in transmission and that a reduction in mask usage impacts the reduction in transmission proportionately.^11^ Therefore, the following scaling factor (due to mask use) was applied daily to the base contact matrix:

$${Scaling Factor}_{Mask use}=1-\left( Mask use\left( \% \right)\times0.30 \right)$$

Figure 4. Change in mask use over time 2020 - 2022

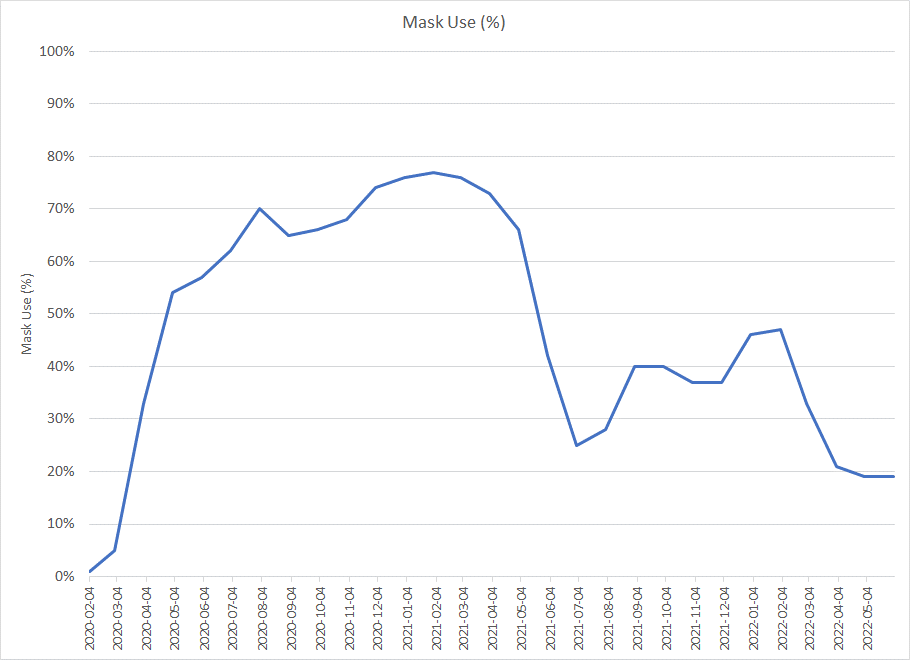

Daily estimates of the change in mobility were obtained from IHME^11^ and was applied to reduce the number of contacts per person. Since age-specific data on changes in mobility were not available, the impact was applied equally to all age groups. Therefore, the following scaling factor (due to change in mobility) was applied daily to the base contact matrix:

$${Scaling Factor}_{Mobility}=1+Change in mobility (\%)$$

The overall scaling factor is thus calculated as:

$$Overall {Scaling Factor}=\left[ {Scaling Factor}_{Mask use} \right] \times\left[ {Scaling Factor}_{Mobility} \right]$$

$$Overall {Scaling Factor}=\left[ 1-\left( Mask use\left( \% \right)\times0.30 \right) \right] \times\left[ 1+Change in mobility (\%) \right]$$

Linear interpolation was used to estimate the daily overall scaling factor based on the monthly estimates.

Figure 5. Change in mobility over time 2020 - 2022

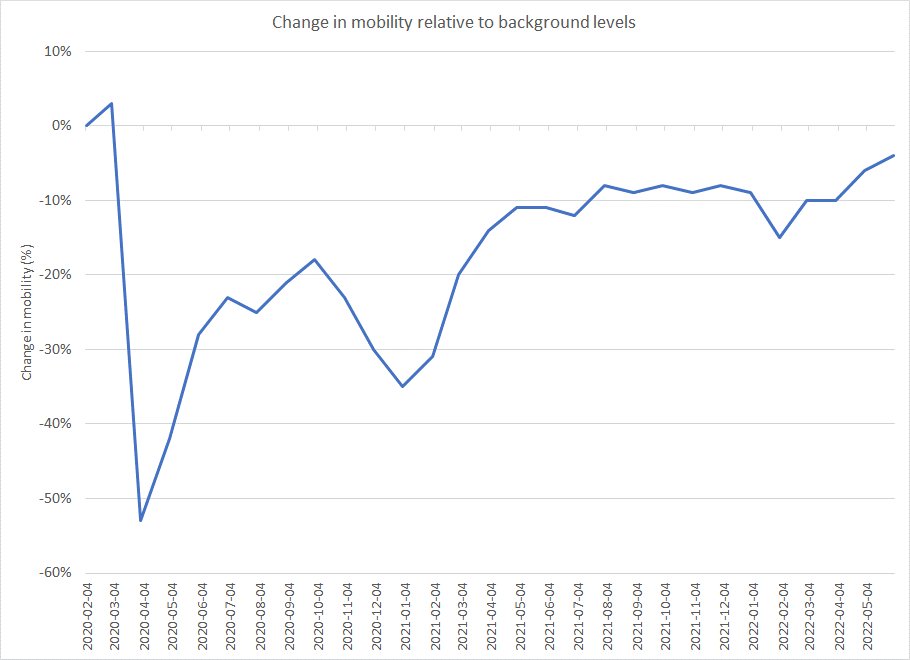

**Vaccine Effectiveness**

Pre-Omicron Period

Initial VE for the primary series of each vaccine against infection and severe disease was obtained from the December 22, 2021 IHME COVID-19 model update.^14^ Monthly waning for each vaccine was approximated from the IHME graphs (timepoint 0-50 weeks), which were based on pooled data from 20 studies that estimated VE as a function of time.^14^ Infection VE for mRNA-1273 and BNT162b2 boosters were obtained from Andrews et al., 2022,^15^ who used a test-negative case-control study to estimate VE in England. The booster infection VE for AD26.COV2.S was assumed by taking the ratio of primary series infection VE for AD26.COV2.S and mRNA-1273, and applying it to the booster infection VE for Moderna.

Booster VE for severe disease for each vaccine was estimated by assuming the same ratio between severe disease and infection observed from the primary series VE for each vaccine and applying it to the booster VE for infection. Monthly waning for booster infection and severe disease was assumed to be the same as primary series values.

Omicron BA1 Period

Primary series initial VEs against infection (all vaccines) and severe disease (BNT162.b2, AD26.COV2.S) were obtained from a meta-analysis conducted by Pratama et al., 2022.^16^ Severe disease VE for mRNA-1273 was obtained from the United Kingdom (UK) Health Security Agency June 16, 2022 COVID-19 vaccine surveillance report.^17^ VE monthly waning values for infection (mRNA-1273, BNT162b2) and severe disease (BNT162b2) were obtained from Pratama et al., 2022.^16^ Monthly waning for severe disease for AD26.COV2.S was unavailable and assumed to be the same as AD26.COV2.S waning against infection.

Booster initial VEs against infection and severe disease for all vaccines, were obtained from Pratama et al., 2022.^16^ Waning against infection and severe disease for boosters were based on smaller sample sizes than primary series. Therefore, booster waning was assumed to be the same as for primary series.

Table 4. Vaccine effectiveness for the pre-Omicron and Omicron (BA.1) periods: by vaccine type

| Vaccine | **Pre-Omicron** | | **Omicron (BA.1)** | |
| --- | --- | --- | --- | --- |
|  | **Initial VE** | **Monthly decline** | **Initial VE** | **Monthly** |
| **Primary Series** | | | | |
| I**nfection** | | | | |
| mRNA-1273 | 91.0% | 3.4% | 76.1% | 9.6% |
| BNT162b2 | 84.0% | 4.0% | 58.4% | 5.9% |
| AD26.COV2.S | 64.0% | 4.0% | 59.9% | 7.0% |
| **Severe Disease** | | | | |
| mRNA-1273 | 97.0% | 1.0% | 90.0% | 3.6% |
| BNT162b2 | 95.0% | 1.3% | 89.9% | 2.7% |
| AD26.COV2.S | 76.0% | 2.1% | 67.0% | 7.0% |
| **Booster** | | | | |
| **Infection** | | | | |
| mRNA-1273 | 95.3% | 3.4% | 78.4% | 9.6% |
| BNT162b2 | 92.3% | 4.0% | 68.8% | 5.9% |
| AD26.COV2.S | 67.0% | 4.0% | 66.0% | 7.0% |
| **Severe Disease** | | | | |
| mRNA-1273 | 100.0% | 1.0% | 99.2% | 3.6% |
| BNT162b2 | 100.0% | 1.3% | 89.1% | 2.7% |
| AD26.COV2.S | 78.4% | 2.1% | 86.0% | 7.0% |

VE, vaccine effectiveness.

The number of vaccines administered were obtained from the CDC COVID Data Tracker for people completing the primary series and first booster.^24^ The proportion of each vaccine administered for the primary series and the first booster are shown in Table 5, with most people receiving an mRNA vaccine. The effectiveness for each vaccine was weighted by these proportions to produce the average vaccine effectiveness.

Table 5. Market share for vaccines delivered for the primary series and first booster in the United States (By June 20, 2022)

|  | **Primary Series**  **(“Fully Vaccinated”)** | **First Booster** |
| --- | --- | --- |
| mRNA-1273 | 34.8% | 42.6% |
| BNT162b2 | 57.5% | 55.9% |
| AD26.COV2.S | 7.7% | 1.5% |

**Transmissibility**

Model calibration was conducted to estimate the transmissibility parameter that reflects cases of COVID-19 experienced in the United States from February 2020 through May 2022. The analytical choices for the calibration process were made considering the recommendations of Vanni and colleagues.^35^

Inputs Varied During Calibration

Similar to the previously develop model by Shiri et al.,^10,36^ the transmissibility parameter, an input in the force of infection equation, was varied during the calibration process. The transmissibility parameter was allowed to vary on a daily basis. For the calibration process, transmissibility parameters were manually varied monthly (days 1, 30, 61, 91, 122, 152, 183, 214, 244, 275, 305, 336, 367, 395, 426, 456, 487, 517, 548, 579, 609, 640, 670, 701, 732, 760, 791) and linear interpolation was assumed between these days (Table 12).

Calibration Targets

The calibration target was the daily incidence of reported COVID-19 cases (symptomatic and asymptomatic), as reported by the IHME during the period of February 4, 2020 through May 31, 2022.

Goodness of Fit Measure

An initial model calibration was performed in a qualitative way by visually comparing the daily incidence of COVID cases predicted by the model and the values obtained from the IHME data.

Results of the Calibration

The final transmissibility parameters for the base case scenario are summarized in Table 6. The comparison plot of the daily number of incident cases of COVID predicted from the calibrated model and the corresponding estimates from the IHME is presented in Figure 6.

Table 6. Final model inputs for the dynamic model

| **Days from Start of Analysis** | **Corresponding Date** | **Transmissibility parameter estimate** |
| --- | --- | --- |
| 31-Jan-2020 | 1 | 0.0238 |
| 29-Feb-2020 | 30 | 0.0286 |
| 31-Mar-2020 | 61 | 0.0343 |
| 30-Apr-2020 | 91 | 0.0190 |
| 31-May-2020 | 122 | 0.0152 |
| 30-Jun-2020 | 152 | 0.0200 |
| 31-Jul-2020 | 183 | 0.0143 |
| 31-Aug-2020 | 214 | 0.0124 |
| 30-Sep-2020 | 244 | 0.0152 |
| 31-Oct-2020 | 275 | 0.0219 |
| 30-Nov-2020 | 305 | 0.0219 |
| 31-Dec-2020 | 336 | 0.0200 |
| 31-Jan-2021 | 367 | 0.0143 |
| 28-Feb-2021 | 395 | 0.0133 |
| 31-Mar-2021 | 426 | 0.0200 |
| 30-Apr-2021 | 456 | 0.0152 |
| 31-May-2021 | 487 | 0.0124 |
| 30-Jun-2021 | 517 | 0.0229 |
| 31-Jul-2021 | 548 | 0.0286 |
| 31-Aug-2021 | 579 | 0.0229 |
| 30-Sep-2021 | 609 | 0.0171 |
| 31-Oct-2021 | 640 | 0.0200 |
| 30-Nov-2021 | 670 | 0.0267 |
| 31-Dec-2021 | 701 | 0.0362 |
| 31-Jan-2022 | 732 | 0.0143 |
| 28-Feb-2022 | 760 | 0.0095 |
| 31-Mar-2022 | 791 | 0.0181 |
| 30-Apr-2022 | 821 | 0.0200 |
| 31-May-2022 | 852 | 0.0133 |

Figure 6. Comparison of daily incidence of COVID cases (calibrated model estimates vs. IHME estimate)

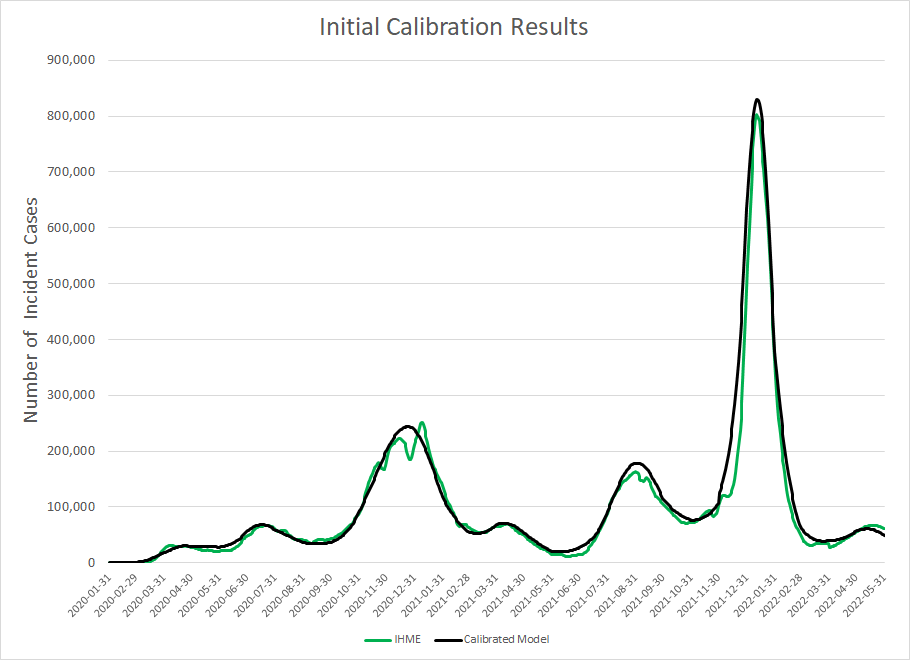

IHME: Institute for Health Metrics and Evaluation

Figure 7. Comparison of daily incidence of COVID cases (calibrated model vs. IHME) with the corresponding transmissibility parameter estimate

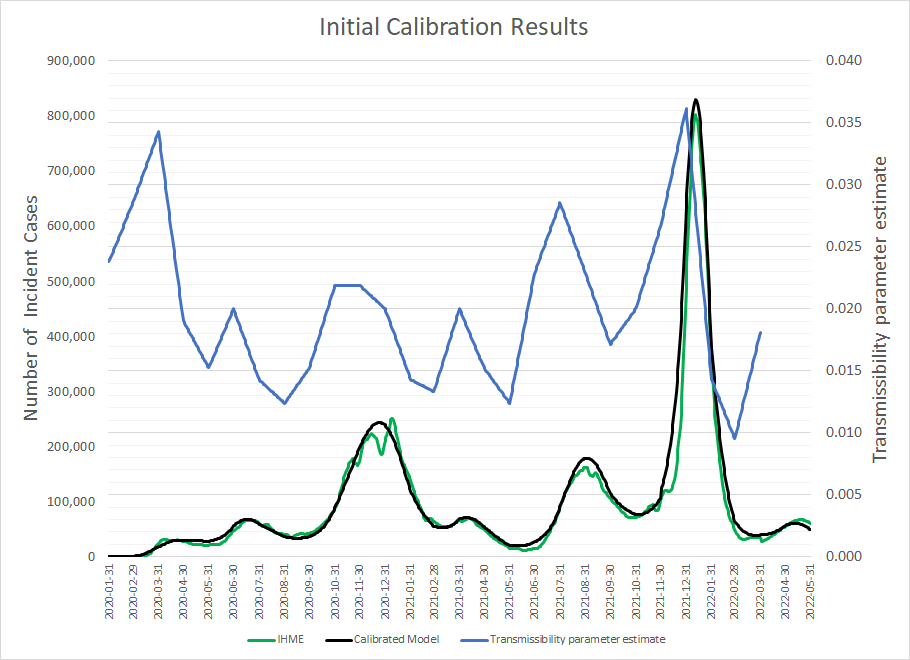

IHME: Institute for Health Metrics and Evaluation

### 1.3. Consequences of infections

#### 1.3.1. Percentage of infections with symptoms

The age-specific proportions of infections that are symptomatic were calculated from Reese et al., (2021)^25^ using data from February to September 2020; laboratory-confirmed case counts that were reported nationally were adjusted for sources of under-detection.**^25^**

Table 7. Percent of infections that are symptomatic

| **Age Group (years)** | **Percentage of Symptomatic Cases*** | **Data Source** |
| --- | --- | --- |
| 18-49 | 85.3% | Reese et al., (2021) **^25^** |
| 50-64 | 85.1% |  |
| ≥65 | 80.8% |  |

*Calculated by dividing the number of symptomatic cases by the total number of infections

#### 1.3.2. Hospitalization Rates

Age-specific hospitalization rates among unvaccinated individuals (Table 8) were calculated from Reese et al., (2021) **^25^** by dividing the total number of estimated hospitalizations (accounting for underreporting) by the total number of symptomatic cases. As Omicron is thought to be less severe than previous VOCs, the rates of hospitalization were reduced using data from Wang et al., 2022, a retrospective cohort study designed to compare severe clinical outcomes between propensity score matched Delta and Omicron cohorts.^27^

Table 8. Age-specific hospitalization rates in unvaccinated people

| **Age Group (years)** | **Pre-Omicron Hospitalization Rates ^25^** | **Omicron Hospitalization Rates** |
| --- | --- | --- |
| 18-49 | 2.6% | 1.25% |
| 50-64 | 7.2% | 3.45% |
| ≥65 | 22.4% | 15.71% |

.

#### 1.3.3. Novel Treatments

The model includes Paxlovid™ (nirmatrelvir and ritonavir), which has been granted emergency use authorization (EUA) by the US Food and Drug Administration (FDA) for the treatment of mild-to-moderate COVID-19 in adults and pediatric patients (12 years of age and older) with positive results of direct SARS-CoV-2 viral testing, and who are at high risk for progression to severe COVID-19, including hospitalization or death.^37^ Paxlovid was selected for inclusion in a sensitivity analysis as it is the National Institute of Health’s preferred treatment, given that it is an oral medication and has the strongest efficacy against hospitalization and death.^38^ Paxlovid is an oral prescription medication that should be taken as soon as possible following diagnosis, within 5 days of symptom onset, and consists of 300 mg nirmatrelvir (two 150 mg tablets) with 100 mg ritonavir (one 100 mg tablet), with all three tablets taken together twice daily for 5 days.^39^

The proportions of patients at high-risk of severe outcomes from COVID-19 infection, and therefore eligible for novel treatments, were obtained from Qasmieh et al., (2022)^28^, along with estimates of Paxlovid utilization. Paxlovid efficacy was assumed to be equivalent in vaccinated and unvaccinated patients.

Table 9. Inputs related to Paxlovid use

| **Model Parameter** | **Value** | **Data Source** |
| --- | --- | --- |
| *Proportion of patients eligible for antivirals**^†^ | | |
| ≥ 18 years | 15.1% | Qasmieh (2022)^28^ |
| *Paxlovid Utilization Rates* | | |
| 18-64 years | 17.2% | Qasmieh (2022)^28^ |
| ≥ 65 years | 2.1% |  |
| *Paxlovid Efficacy* | | |
| Reduced risk of hospitalization or death | 88% | EPIC-HR Trial^40^ |

*Eligibility defined as ≥65 years or with comorbidities (cancer, diabetes, obesity, chronic obstructive pulmonary disease or lung disease, liver disease, heart disease, high blood pressure, a recent organ transplant, or an immunodeficiency), with reported symptoms and tested positive on an at-home rapid or point of care rapid or PCR test

### 1.4. Equations associated with the SEIR Model

#### 1.4.1. Differential Equations

**Unvaccinated cohort**

$$S_{t+1,j}^{X}=S_{t,j}^{X}-\lambda_{t,j}^{X}S_{t,j}^{X}-\mu_{t,j}+\omega_{t,j}^{X}R_{t,j}^{X}-\epsilon_{t,j}$$

$$E_{t+1,j}^{X}=E_{t,j}^{X}+\lambda_{t,j}^{X}S_{t,j}^{X}-\frac{1}{\tau_{E}}E_{t,j}^{X}$$

$$I_{t+1,j}^{X}=I_{t,j}^{X}+\frac{1}{\tau_{E}}E_{t,j}^{X}-\frac{1}{\tau_{I}}I_{t,j}^{X}+\epsilon_{t,j}$$

$$R_{t+1,j}^{X}=R_{t,j}^{X}+\frac{1}{\tau_{I}}I_{t,j}^{X}-\omega_{t,j}^{X}R_{t,j}^{X}$$

**Vaccinated cohort**

$$S_{t+1,j}^{V}=S_{t,j}^{V}-\lambda_{t,j}^{V}S_{t,j}^{V}+\mu_{t,j}-\nu_{t,j}-{\left( p_{t,j}^{V,B3} \right)\nu}_{t,j}^{3}+\omega_{t,j}^{V}R_{t,j}^{V}$$

$$E_{t+1,j}^{V}=E_{t,j}^{V}+\lambda_{t,j}^{V}S_{t,j}^{V}-\frac{1}{\tau_{E}}E_{t,j}^{V}$$

$$I_{t+1,j}^{V}=I_{t,j}^{V}+\frac{1}{\tau_{E}}E_{t,j}^{V}-\frac{1}{\tau_{I}}I_{t,j}^{V}$$

$$R_{t+1,j}^{V}=R_{t,j}^{V}+\frac{1}{\tau_{I}}I_{t,j}^{V}-\omega_{t,j}^{V}R_{t,j}^{V}$$

**Boosted cohort**

$$S_{t+1,j}^{B}=S_{t,j}^{B}-\lambda_{t,j}^{B}S_{t,j}^{B}+\nu_{t,j}-\nu_{t,j}^{2}-{\left( p_{t,j}^{B,B3} \right)\nu}_{t,j}^{3}+\omega_{t,j}^{B}R_{t,j}^{B}$$

$$E_{t+1,j}^{B}=E_{t,j}^{B}+\lambda_{t,j}^{B}S_{t,j}^{B}-\frac{1}{\tau_{E}}E_{t,j}^{B}$$

$$I_{t+1,j}^{B}=I_{t,j}^{B}+\frac{1}{\tau_{E}}E_{t,j}^{B}-\frac{1}{\tau_{I}}I_{t,j}^{B}$$

$$R_{t+1,j}^{B}=R_{t,j}^{B}+\frac{1}{\tau_{I}}I_{t,j}^{B}-\omega_{t,j}^{B}R_{t,j}^{B}$$

**Boosted cohort (Second dose)**

$$S_{t+1,j}^{B2}=S_{t,j}^{B2}-\lambda_{t,j}^{B2}S_{t,j}^{B2}+\nu_{t,j}^{2}-{\left( p_{t,j}^{B2,B3} \right)\nu}_{t,j}^{3}+\omega_{t,j}^{B2}R_{t,j}^{B2}$$

$$E_{t+1,j}^{B2}=E_{t,j}^{B2}+\lambda_{t,j}^{B2}S_{t,j}^{B2}-\frac{1}{\tau_{E}}E_{t,j}^{B2}$$

$$I_{t+1,j}^{B2}=I_{t,j}^{B2}+\frac{1}{\tau_{E}}E_{t,j}^{B2}-\frac{1}{\tau_{I}}I_{t,j}^{B2}$$

$$R_{t+1,j}^{B2}=R_{t,j}^{B2}+\frac{1}{\tau_{I}}I_{t,j}^{B2}-\omega_{t,j}^{B2}R_{t,j}^{B2}$$

**Boosted cohort (Third dose)**

$$S_{t+1,j}^{B3}=S_{t,j}^{B3}-\lambda_{t,j}^{B3}S_{t,j}^{B3}+\left( p_{t,j}^{V,B3}+p_{t,j}^{B,B3}+p_{t,j}^{B2,B3} \right)\nu_{t,j}^{3}+\omega_{t,j}^{B3}R_{t,j}^{B3}$$

$$E_{t+1,j}^{B3}=E_{t,j}^{B3}+\lambda_{t,j}^{B3}S_{t,j}^{B3}-\frac{1}{\tau_{E}}E_{t,j}^{B3}$$

$$I_{t+1,j}^{B3}=I_{t,j}^{B3}+\frac{1}{\tau_{E}}E_{t,j}^{B3}-\frac{1}{\tau_{I}}I_{t,j}^{B3}$$

$$R_{t+1,j}^{B3}=R_{t,j}^{B3}+\frac{1}{\tau_{I}}I_{t,j}^{B3}-\omega_{t,j}^{B2}R_{t,j}^{B3}$$

**For t=1,**

$S_{1,j}^{X}$ = Initial number of susceptible individuals

$I_{1,j}^{X}$= Initial number of infectious individuals = $\epsilon_{1,j}$

$$E_{1,j}^{X}=R_{1,j}^{X}=0$$

$$S_{1,j}^{V}=E_{1,j}^{V}=I_{1,j}^{V}=R_{1,j}^{V}=0$$

$$S_{1,j}^{B}=E_{1,j}^{B}=I_{1,j}^{B}=R_{1,j}^{B}=0$$

$$S_{1,j}^{B2}=E_{1,j}^{B2}=I_{1,j}^{B2}=R_{1,j}^{B2}=0$$

$$S_{1,j}^{B3}=E_{1,j}^{B3}=I_{1,j}^{B3}=R_{1,j}^{B3}=0$$

Notes, superscripts:

*X*, *V*, *B*, *B2*, and *B3* represent the unvaccinated, vaccinated, boosted, boosted (second dose), and boosted (third dose) cohorts, respectively.

Notes, subscripts:

*t* = time (i.e., day of analysis)

*j* = age group (number 1 to 9)

**Definitions**

$S_{j}^{X}, S_{j}^{V}, S_{j}^{B}, S_{j}^{B2},S_{j}^{B3}$ represent the proportion of susceptibles in age group *j* in cohort *X*, *V, B,* *B2*, or *B3*.

The compartments with superscript *X*, *V*, and *B* represent the unvaccinated, vaccinated, and boosted cohorts, respectively.

$E_{j}^{Z}$ represent exposed, but not yet infectious, individuals in age group *j* in cohort *Z* (*X*, *V*, *B,* *B2*, or *B3*).

$I_{j}^{Z}$ represent infectious individuals in age group *j* in cohort *Z* (*X*, *V*, *B,* *B2*, or *B3*).

$R_{j}^{Z}$ represent immune individuals in age group *j* in cohort *Z* (*X*, *V*, *B,* *B2*, or *B3*).

$\lambda_{t,j}^{*}$is the age-group specific force of infection (see section below)

$\frac{1}{\tau_{E}}$ is the rate of loss of latency

$\frac{1}{\tau_{I}}$ is the rate of loss of infectiousness

$\mu_{t,j}$ is the proportion receiving a vaccination on day *t* in age group *j*

$\nu_{t,j}$ is the proportion receiving a booster on day *t* in age group *j*

$\nu_{t,j}^{2}$ is the proportion receiving a second booster on day *t* in age group *j*

$\nu_{t,j}^{3}$ is the proportion receiving a third booster on day *t* in age group *j*

$p_{t,j}^{V,B3}$ is the proportion receiving a third booster from cohort *V* on day *t* in age group *j*

$p_{t,j}^{B,B3}$ is the proportion receiving a third booster from cohort *B* on day *t* in age group *j*

$p_{t,j}^{B2,B3}$ is the proportion receiving a third booster from cohort *B2* on day *t* in age group *j*

Note: $p_{t,j}^{V,B3}+p_{t,j}^{B,B3}+p_{t,j}^{B2,B3}=1$

$\omega_{t,j}^{X}$ is the natural immunity waning rate on day *t* in age group *j* in cohort *X*, *V*, *B,* *B2*, or *B3*

$\epsilon_{t,j}$ is the proportion of external cases on day *t* in age group *j*

#### 1.4.2. Force of Infection (Unvaccinated)

$$\lambda_{t,i}^{*}=\left( {Scaling Factor}_{Mask use, t} \right)\times\left( {Scaling Factor}_{Mobility,t} \right)\times\left[ \beta_{t}\sum_{j=1}^{9} \sum_{Z=\left\{ X,V,B \right\}} c_{ij}I_{t,j}^{Z} \right]$$

$\lambda_{t,i}^{*}$is the age-group specific force of infection at time *t* for age group *i*

${Scaling Factor}_{Mask use, t}$at time *t* is defined in section 0

${Scaling Factor}_{Mobility,t}$ at time *t* is defined in section 0

$\beta_{t}$ is the transmissibility parameter at time *t*

$c_{ij}$ is the rate at which individuals in age group *i* make contact with those in age group *j*

$I_{t,j}^{Z}$ represents the infectious individuals in cohort *Z* at time *t* for age group *j*.

#### 1.4.3. Force of Infection (Vaccinated)

For the vaccinated cohorts, the force of infection calculation is adjusted based on the vaccine effectiveness in the cohort:

$$\lambda_{t,i}^{Z}=\left( 1-{VE}_{t,i}^{X} \right)\times\left( {Scaling Factor}_{Mask use, t} \right)\times\left( {Scaling Factor}_{Mobility,t} \right)\times\left[ \beta_{t}\sum_{j=1}^{9} \sum_{Z=\left\{ X,V,B \right\}} c_{ij}I_{t,j}^{Z} \right]$$

$\lambda_{t,i}^{Z}$is the age-group specific force of infection at time *t* for age group *i* in cohort *Z* (*X*, *V*, *B,* *B2*, or *B3*).

${VE}_{t,i}^{X}$ is the vaccine effectiveness at time *t* for age group *i* in cohort *Z* (*X*, *V*, *B,* *B2*, or *B3*).
